# Trans-ancestral genetic study of diabetes mellitus risk in survivors of childhood cancer: a report from the St. Jude Lifetime Cohort and the Childhood Cancer Survivor Study

**DOI:** 10.1101/2023.06.02.23290868

**Authors:** Cindy Im, Achal Neupane, Jessica L. Baedke, Angela Delaney, Stephanie B. Dixon, Eric J. Chow, Sogol Mostoufi-Moab, Melissa A. Richard, M. Monica Gramatges, Philip J. Lupo, Noha Sharafeldin, Smita Bhatia, Gregory T. Armstrong, Melissa M. Hudson, Kirsten K. Ness, Leslie L. Robison, Yutaka Yasui, Carmen L. Wilson, Yadav Sapkota

## Abstract

Type 2 diabetes mellitus (T2D) is an established late effect of treatment for childhood cancer. Leveraging detailed cancer treatment and whole-genome sequencing data among survivors of childhood cancer of European (EUR) and African (AFR) genetic ancestry in the St. Jude Lifetime Cohort (N=3,676; 304 cases), five novel diabetes mellitus (DM) risk loci were identified with independent trans-/within-ancestry replication, including in 5,965 survivors of the Childhood Cancer Survivor Study. Among these, common risk variants at 5p15.2 (*LINC02112*), 2p25.3 (*MYT1L*), and 19p12 (*ZNF492*) modified alkylating agent-related risks across ancestry groups, but AFR survivors with risk alleles experienced disproportionately greater risk of DM (AFR, variant ORs: 3.95-17.81; EUR, variant ORs: 2.37-3.32). Novel risk locus *XNDC1N* was identified in the first genome-wide DM rare variant burden association analysis in survivors (OR=8.65, 95% CI: 3.02-24.74, P=8.1×10^-6^). Lastly, a general-population 338-variant multi-ancestry T2D polygenic risk score was informative for DM risk in AFR survivors, and showed elevated DM odds after alkylating agent exposures (quintiles: combined OR_EUR_=8.43, P=1.1×10^-8^; OR_AFR_=13.85, P=0.033). This study supports future precision diabetes surveillance/survivorship care for all childhood cancer survivors, including those with AFR ancestry.

---

With advances in therapy, five-year survival after childhood cancer diagnosis has improved to over 85%^1^. However, as a consequence of exposures to these curative therapies, long-term (≥5 years) survivors face excess chronic disease morbidity^2–4^, including increased risk for developing type 2 diabetes mellitus (T2D)^5, 6^ later in life. Survivors demonstrate an almost 3- fold greater risk for diabetes mellitus (DM), primarily T2D^5, 6^, compared to siblings^5, 7^. The risk for DM is elevated among survivors who received abdominal radiation therapy (RT) or total body irradiation, and to a lesser extent, alkylating agents^5, 6, 8, 9^. Additionally, younger ages at primary cancer diagnosis, increasing age, and higher body mass index (BMI) also contribute to risk^6, 9^.

Known clinical risk factors do not fully explain why some survivors develop DM while others do not. Genetic susceptibility is posited to play a role. Genome-wide association studies (GWAS), including large-scale meta-analyses with >1 million participants in the general population, have identified >550 loci associated with T2D risk, including those shared across diverse ancestral groups^10, 11^. Analyses of rare variants (minor allele frequencies <1%) have revealed additional T2D risk loci^12^ and have been shown to enhance T2D risk stratification^13^. But given that DM genetic susceptibility among survivors is anticipated to additionally reflect the risk contributions conferred by cancer treatments, novel investigations of DM genetic risk are also needed among survivors of childhood cancer^14^. A recent GWAS of DM risk among 5,083 survivors of European ancestry participating in the Childhood Cancer Survivor Study (CCSS) in which cases were identified through self-report identified one novel genome-wide significant risk locus at 9q22.32 (*ERCC6L2*)^15^.

To date, our understanding of DM genetic susceptibility factors among childhood cancer survivors are limited while studies including survivors of diverse ancestry or exploring the contribution of rare variants to DM risk do not exist, to our knowledge. Hence, we report findings from a trans-ancestry GWAS and a gene-based rare variant burden analysis with whole-genome sequencing (WGS) data and clinically-ascertained DM status in 3,676 survivors of European and African genetic ancestry participating in the St. Jude Lifetime Cohort (SJLIFE). With the identification of five novel DM risk loci, we highlight the need to study common and rare genetic susceptibility factors contributing to risk of late effects among diverse populations of childhood cancer survivors to provide future precision surveillance/survivorship care.

## RESULTS

The demographic and clinical characteristics of SJLIFE childhood cancer survivors included in trans-ancestral and rare variant discovery analyses are provided in Table 1. A total of 261 (8.4%) and 43 (7.5%) survivors of European genetic ancestry (EUR; N=3,102) and African genetic ancestry (AFR; N=574) developed DM, respectively. SJLIFE AFR survivors were comparatively younger at follow up (median=28 years, IQR: 21-37 years) than SJLIFE EUR survivors (median=32 years, IQR: 24-41 years). The prevalence of overweight/obesity (BMI >25 kg/m^2^) was similar between ancestry groups (∼60%). Nearly one-fifth of SJLIFE survivors received abdominal RT, while 57% and 52% of EUR and AFR survivors were treated with alkylating agents, respectively. Non-genetic covariates used in downstream genetic analyses were the same as those used in a previous DM GWAS in CCSS^15^, with abdominal RT and alkylating agents as cancer treatment risk factors of interest (Supplemental Table 1). BMI was significantly associated with DM risk in SJLIFE survivors (EUR: OR=1.56 per 5 kg/m^2^, P=3.8×10^-28^; AFR: OR=1.45 per 5 kg/m^2^, P=3.9×10^-4^), followed by abdominal RT dose (OR=1.23 per 10 Gray, P=3.6×10^-4^) and any treatment with alkylating agents (OR=1.35, 95% CI: 1.02-1.80, P=0.039) among EUR survivors. For the replication study conducted in EUR survivors participating in the Childhood Cancer Survivor Study (CCSS), 223 (5.7%) and 47 (2.3%) self-reported use of glucose-lowering medications to treat DM in the Original (cancer diagnosed 1970-1986) and Expansion (cancer diagnosed 1986-1999) cohorts, respectively (Supplementary Table 2).

**Table 1:** Demographic and clinical characteristics of childhood cancer survivors in the SJLIFE cohort, by ancestry

| Characteristic | SJLIFE, European ancestry<br>(N=3102) |  | SJLIFE African ancestry<br>(N=574) |  |
| --- | --- | --- | --- | --- |
|  | N | % | N | % |
| Sex |  |  |  |  |
| Male | 1648 | 53.1 | 286 | 49.8 |
| Female | 1454 | 46.9 | 288 | 50.2 |
| Age at follow up (in years) |  |  |  |  |
| <25 | 904 | 29.1 | 228 | 39.7 |
| 25-34 | 957 | 30.9 | 179 | 31.2 |
| 35-44 | 731 | 23.6 | 107 | 18.6 |
| ≥45 | 510 | 16.4 | 60 | 10.5 |
| Body mass index<sup>a</sup> (kg/m<sup>2</sup>) |  |  |  |  |
| ≤25 | 1159 | 37.4 | 217 | 37.8 |
| >25 to ≤30 | 822 | 26.5 | 133 | 23.2 |
| >30 to ≤40 | 899 | 29.0 | 163 | 28.4 |
| >40 | 222 | 7.2 | 61 | 10.6 |
| Developed type 2 diabetes | 261 | 8.4 | 43 | 7.5 |
| Age at cancer diagnosis (in years) |  |  |  |  |
| <5 | 1323 | 42.7 | 244 | 42.5 |
| 5-9 | 672 | 21.7 | 119 | 20.7 |
| 10-14 | 630 | 20.3 | 129 | 22.5 |
| ≥15 | 477 | 15.4 | 82 | 14.3 |
| Childhood cancer diagnosis |  |  |  |  |
| Leukemias | 1040 | 33.5 | 115 | 20.0 |
| Central nervous system tumors | 436 | 14.1 | 81 | 14.1 |
| Hodgkin lymphoma | 343 | 11.1 | 60 | 10.5 |
| Non-Hodgkin lymphoma | 203 | 6.5 | 43 | 7.5 |
| Kidney tumors | 184 | 5.9 | 62 | 10.8 |
| Soft tissue sarcomas | 193 | 6.2 | 52 | 9.1 |
| Other | 703 | 22.7 | 161 | 28.1 |
| Cancer treatments |  |  |  |  |
| Any abdominal radiation therapy (RT) | 566 | 18.3 | 106 | 18.5 |
| Any alkylating agents | 1751 | 56.5 | 299 | 52.1 |
| Dosages: Median (IQR)<sup>b</sup> |  |  |  |  |
| Abdominal RT tumor dose (cGy) | 566 | 2400 (2000-3500) | 106 | 2050 (1200-3375) |
| Alkylating agents (CED, mg/m<sup>2</sup>) | 1751 | 8155 (4425-13271) | 299 | 7448 (3696-11783) |

Abbreviations: SJLIFE, St. Jude Lifetime Cohort; IQR, interquartile range; RT, radiation therapy; CED, cyclophosphamide equivalent dose; mg, milligrams; m, meters; kg, kilograms.
a. Body mass index values were adjusted for amputation.
b. Treatment dose distribution statistics are provided for childhood cancer survivors who received any of the specified treatments and had non-missing dosage information.

### Common genetic variants with trans-ancestry associations with DM risk

We conducted ancestry-specific GWAS for clinically-ascertained DM in SJLIFE with logistic regression models adjusted for relevant clinical and sociodemographic risk factors, separately analyzing 3,102 EUR and 574 AFR survivors. To facilitate discovery of common loci with trans-ancestral DM risk effects in survivors, we prioritized all variants within a 100-kb window of the most significantly associated index variant (P<5×10^-6^) with effect allele frequencies (EAFs) ≥1% in both SJLIFE EUR and AFR samples for further investigation, using trans-ancestral replication (i.e., replication between ancestry groups, with a Bonferroni-corrected p-value threshold) and trans-ancestry meta-analysis to declare genome-wide significance (P<5×10^-8^) (see Methods). We then utilized available data from CCSS EUR survivors for replication of GWAS findings that were genome-wide significant in the EUR-specific GWAS, and for secondary replication of genome-wide significant trans-ancestry meta-analysis findings. Replication in a second sample of AFR survivors was not feasible in CCSS due to sample size limitations (N=149, with 6 cases).

Genetic variants with replicated trans-ancestral and ancestry-specific DM risk effects are shown in Table 2 (Manhattan and quantile-quantile plots from SJLIFE ancestry-specific GWAS provided in Supplementary Figures 1 and 2). In the EUR-specific GWAS in SJLIFE, we identified one genome-wide significant locus on 8q11.21 within *SNTG1* (syntrophin-gamma 1) (Supplementary Figure 1). The most significantly associated variant at 8q11.21 in the SJLIFE EUR discovery dataset was rs6993752 (OR=1.99, 95% CI: 1.56-2.55; P=4.4×10^-8^) (Table 2, Supplementary Table 3), replicating in CCSS EUR survivors (OR=1.56, 95% CI: 1.12-2.17; P=8.1×10^-3^) (Table 2, Supplementary Tables 4, 5). In SJLIFE AFR survivors, these 8q11.21 variants were associated with DM (P<0.05) but showed ancestral allelic effect heterogeneity (Supplementary Table 3), which can occur due to differences in LD with causal variants, environment, or polygenic background across ancestrally diverse populations^16^. Effect allele frequencies for 8q11.21 variants were generally higher in SJLIFE AFR survivors, but showed a broken pattern of LD in the AFR 1000 Genomes reference panel relative to rs6993752 which was not observed in EUR samples (Supplementary Figure 4).

**Figure 1:**
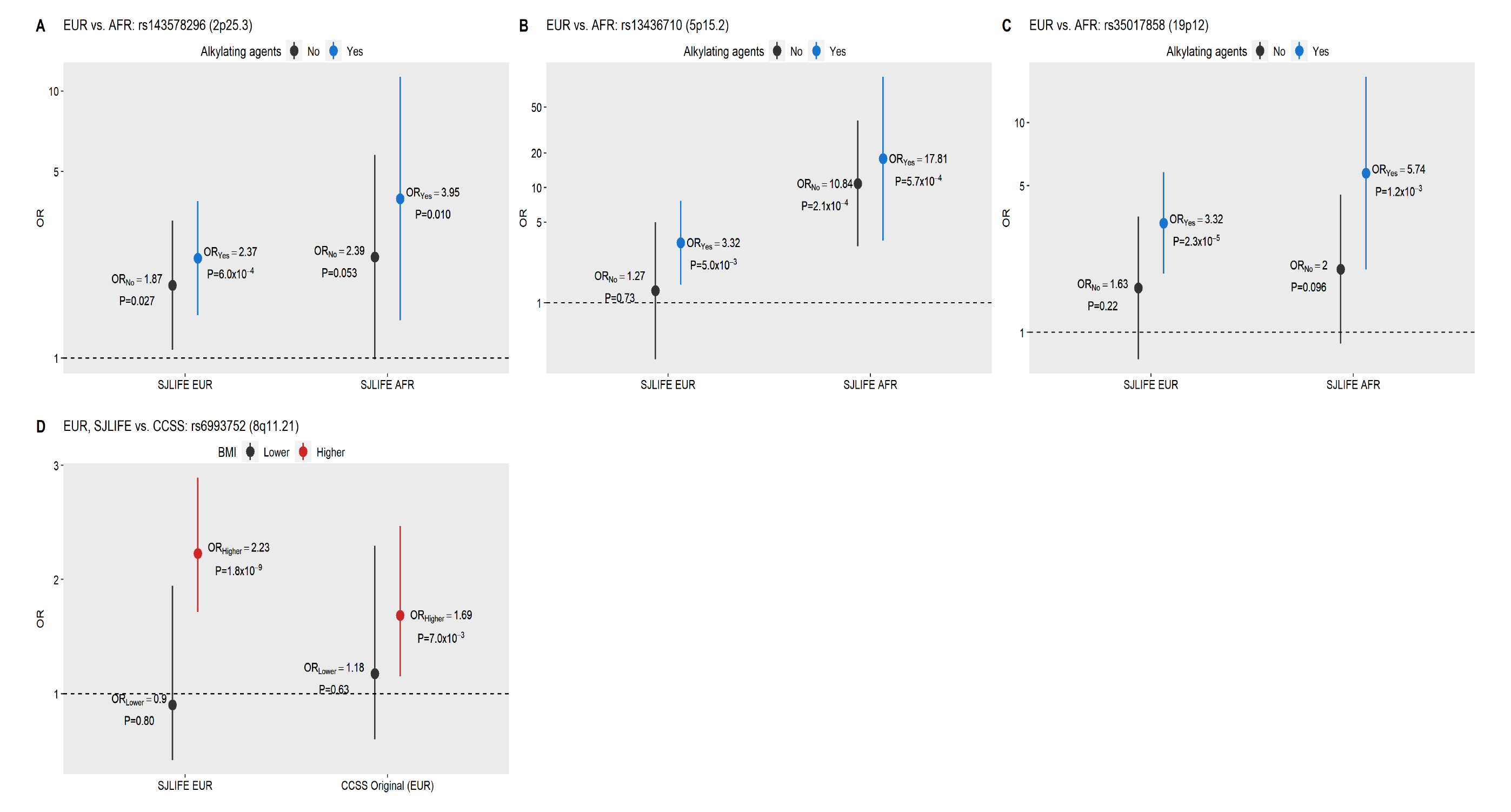
Diabetes mellitus risk associations for replicated index SNPs, stratified by previous treatment with alkylating agents and adiposity. Each panel shows stratified odds ratios (circles) and 95% confidence intervals (whiskers) for the study samples described on the x-axis. Panels A-C show diabetes mellitus risk associations for the risk alleles at the 2p25.3, 5p15.2, and 19p12 loci, respectively, stratified by treatment with alkylating agents (unexposed: black; exposed: blue) in SJLIFE ancestry samples (EUR, European ancestry; AFR, African ancestry). For panels A and C, alkylating agent doses ≥4,000 mg/m^2^ versus none are contrasted, while for panel B, any versus no exposure to alkylating agents are shown (due to lower effect allele frequencies). Panel D shows associations between the risk allele at the 8q11.21 locus (rs6993752) in EUR survivors in SJLIFE and CCSS, stratified by lower body mass index (BMI≤25 kg/m^2^, in black) and higher BMI (>25 kg/m^2^, highlighted in red).

**Figure 2:**
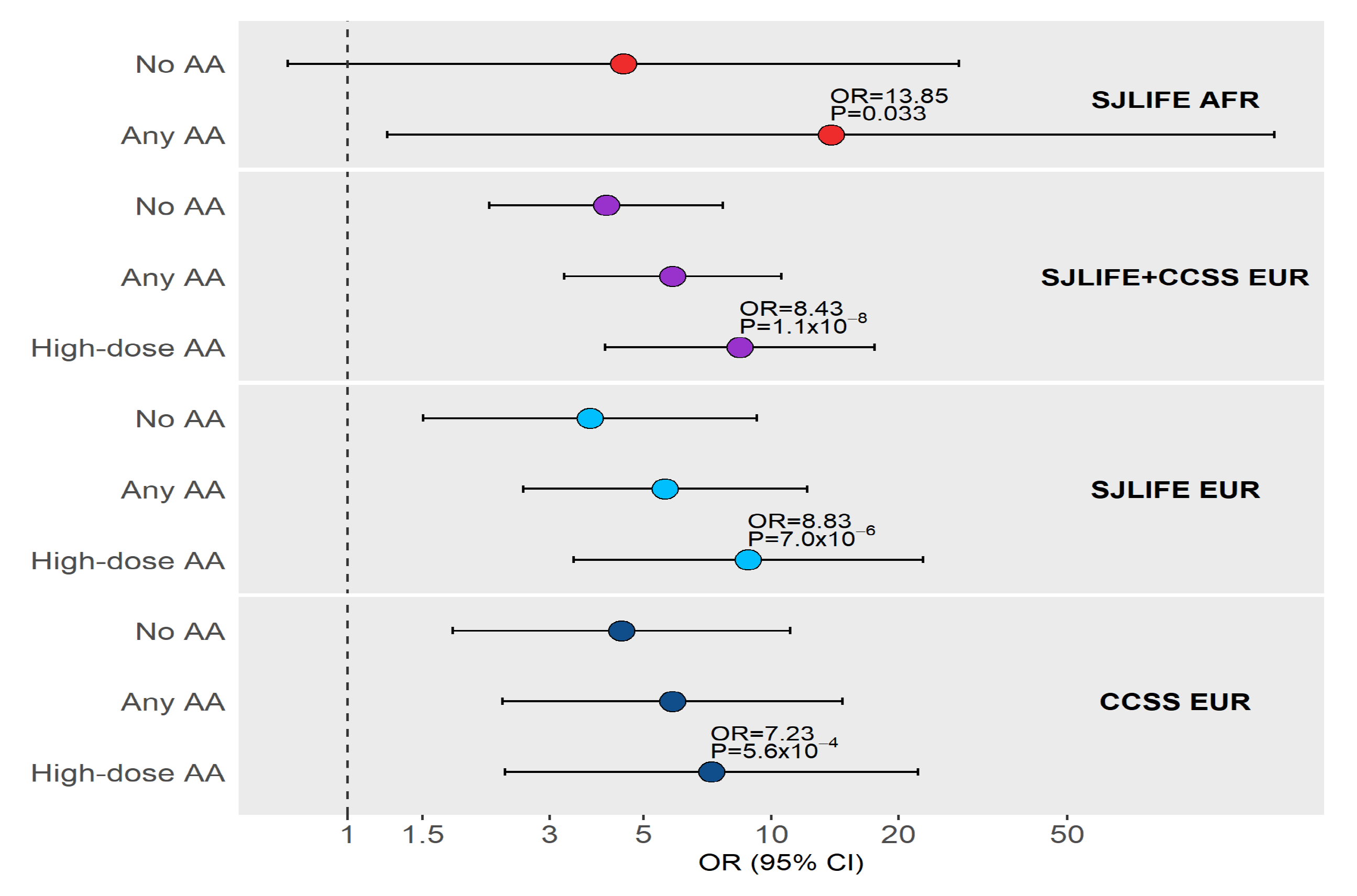
Multi-ancestry PRS quintile associations with diabetes mellitus odds in childhood cancer survivor cohort subgroups defined by ancestry and treatment with alkylating agents (AA). Associations between the top quintile (relative to the lowest quintile) of a general population multi-ancestry PRS for type 2 diabetes (Mahajan *et al.*) and odds of diabetes mellitus in SJLIFE AFR survivors (red), the combined SJLIFE and CCSS cohort of EUR survivors (purple), SJLIFE EUR survivors (light blue), and the combined CCSS EUR survivor cohorts (dark blue) are shown (odds ratios or ORs as circles and corresponding 95% confidence intervals as whiskers). Top PRS quintile ORs and corresponding p-values are annotated for SJLIFE AFR survivors exposed to any AA and for EUR survivors not exposed to abdominal radiation therapy who were treated with high-dose AA (≥4000 mg/m^2^).

**Table 2:**
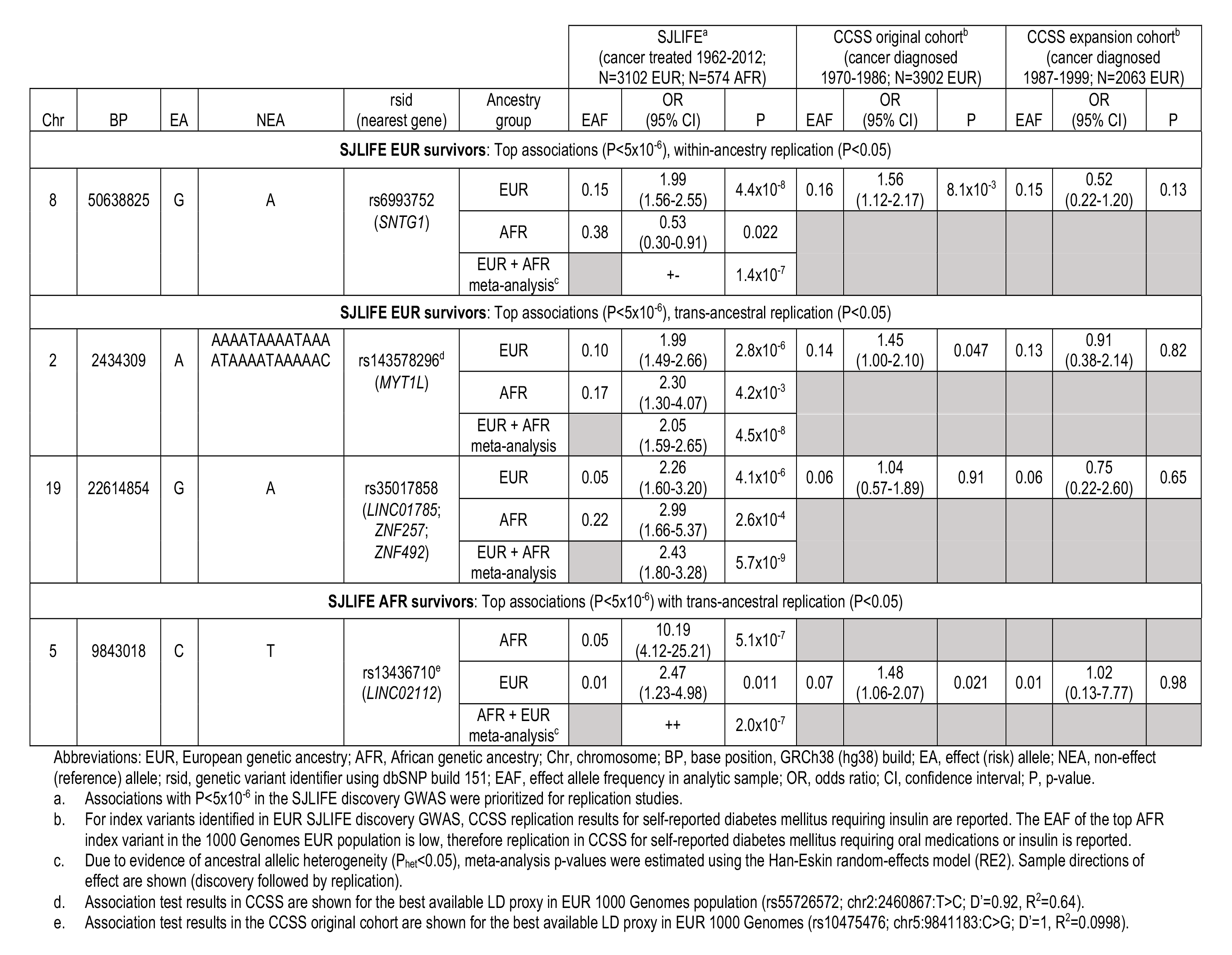
Ancestry-specific and trans-ancestral diabetes risk variants in childhood cancer survivors

We also prioritized two loci with suggestive associations with DM risk (P<5×10^-6^) identified among SJLIFE EUR survivors: 2p25.3, represented by three variants upstream of *MYT1L* (myelin transcription factor 1 like; most significantly associated [index] variant rs143578296: OR=1.99, 95% CI: 1.49-2.66; P=2.8×10^-6^) and 19p12, represented by five variants upstream of *ZNF492* (zinc finger protein 492; index variant rs35017858: OR=2.26, 95% CI: 1.60-3.20; P=4.1×10^-6^) (Table 2, Figure 1). We found that all variants at the 2p25.3 and 19p12 loci were replicated in SJLIFE AFR survivors (Supplementary Table 3) at nominal significance (P<0.05), while the locus index SNPs were replicated under a Bonferroni-corrected p-value threshold that accounted for the number of tested SNPs. The index SNPs at 2p25.3 and 19p12 attained genome-wide significance in trans-ancestry meta-analysis (2p25.3, rs143578296: OR=2.05, 95% CI: 1.59-2.65, P=4.5×10^-8^; 19p12, rs35017858: OR=2.43, 95% CI: 1.80-3.28; P=5.7×10^-9^). Variants at the 2p25.3 locus were also replicated in CCSS EUR survivors (rs55726572 as the best LD proxy, OR=1.45, 95% CI: 1.00-2.10, P=0.047).

In the AFR-specific GWAS in SJLIFE, we identified a novel variant at the 5p15.2 locus (Supplementary Figure 5) with suggestive significance (rs13436710: OR=10.19, 95% CI: 4.12- 25.21, P=5.1×10^-7^). While variants in LD with rs13436710 were low-frequency in EUR samples (EAF ∼1%), index/proxy variants replicated in both SJLIFE EUR (OR=2.47, 95% CI: 1.23-4.98, P=0.011) and CCSS EUR (OR=1.48, 95% CI: 1.06-2.07, P=0.021) survivors.

We examined associations between all available replicated common index variants and self-reported and ICD-coded T2D that were tested in the UK Biobank Study (UKBB; N>361,000) (Supplementary Table 6). The T2D risk effect sizes in the UKBB were extremely modest (OR<1.002) for the index variant at the 2p25.3 locus (rs143578296), which showed nominal association with ICD-10-coded T2D risk (P=3.6×10^-3^), and the index variant at the 19p12 locus (rs35017858), which showed nominal association with self-reported T2D risk (P=6.6×10^-3^).

### Allelic risks for developing DM and the contributions of cancer treatments and adiposity

We investigated whether replicated DM risk variants from common variant analyses in survivors potentially modified DM risk effects following exposures to specific cancer treatments. While risk alleles at the 2p25.3, 5p15.2, and 19p12 loci did not appear to modify abdominal RT-related DM risk effects in both SJLIFE EUR and AFR survivors (Supplementary Table 7), index variants at these loci were associated with appreciably greater DM risks among SJLIFE EUR and AFR survivors treated with alkylating agents, i.e., alkylating agent-related risk effect modifications seen in the discovery data were replicated in another ancestral dataset (Figure 1, Supplementary Table 8). The magnitude of DM associations in SJLIFE AFR survivors treated with alkylating agents was greater for all variants compared to those seen in SJLIFE EUR survivors treated with alkylating agents. For example, the 19p12 index DM risk allele (rs35017858) identified in SJLIFE EUR survivors was associated with 3.3-fold greater DM odds in SJLIFE EUR survivors with high-dose alkylating agent exposures (95% CI: 1.91-5.79, P=2.3×10^-5^), but 5.7-fold greater DM odds in the replication dataset of SJLIFE AFR survivors with high-dose alkylating agent exposures (95% CI: 1.99-16.54, P=1.2×10^-3^). Corresponding allelic risks among survivors without this treatment exposure was appreciably lower (EUR: OR=1.63, P=0.22; AFR: OR=2.00, P=0.096). From the SJLIFE AFR discovery GWAS, the 5p15.2 index variant (rs13436710) risk allele was associated with 17.8-fold (95% CI: 3.46-91.60, P=5.7×10^-4^) greater odds of DM in alkylating agent-exposed AFR survivors, in contrast to the 3.3-fold (95% CI: 1.44-7.68, P=5.0×10^-3^) greater DM odds observed in the replication dataset of SJLIFE EUR survivors. In comparison, corresponding allelic risks among survivors without this treatment exposure were lower (EUR: OR=1.27, P=0.73; AFR: OR=10.84, P=2.1×10^-4^).

The index variant at the 8q11.21 locus (rs6993752) did not show evidence of modifying DM risks related to abdominal RT or alkylating agents. Given treatment with abdominal RT has been previously linked to lower adiposity among survivors^17^ but obesity is strongly associated with other cancer treatments and DM risk^6^, we evaluated whether this DM risk allele showed significant effect heterogeneity by adiposity. Overweight/obese (BMI>25 kg/m^2^) EUR survivors in SJLIFE were significantly more likely to have received cranial RT (P<0.001) or corticosteroids (P<0.001). We found that DM risks associated with the risk allele at the 8q11.21 locus index variant (rs6993752) were appreciably greater among overweight/obese EUR survivors in SJLIFE (OR_BMI>25_=2.23, 95% CI: 1.72-2.89, P=1.8×10^-9^ versus OR_BMI≤25_=0.90, P=0.80) and CCSS (OR_BMI>25_=1.69, 95% CI: 1.15-2.47, P=7.0×10^-3^ versus OR_BMI≤25_=1.18, P=0.63) (Figure 1, Supplementary Table 9).

### T2D polygenic risk in survivors

We assessed whether T2D PRSs derived in non-cancer study populations were associated with DM risk in childhood cancer survivors, and in particular, whether a 338-variant T2D PRS based on general population multi-ancestry GWAS meta-analyses^11^ also had increased transferability in survivors of AFR ancestry. We found standard deviation increases in the multi-ancestry T2D PRS were significantly associated with increased odds of DM in all SJLIFE and CCSS survivor cohorts (SJLIFE AFR: OR=1.80, P=2.8×10^-3^; SJLIFE/CCSS EUR: ORs=1.57-1.84, P<9.5×10^-4^) (Table 3). While a ∼6.9 million-variant PRS derived from a T2D GWAS in Europeans^18^ was also associated with DM in all SJLIFE and CCSS EUR survivor cohorts (ORs: 2.16-3.06, P<6.8×10^-4^), this EUR-only PRS was not associated with DM in SJLIFE AFR survivors (OR=0.97, P=0.95).

**Table 3:** Evaluation of European ancestry-only versus multi-ancestry type 2 diabetes (T2D) polygenic risk scores among childhood cancer survivors in SJLIFE and CCSS

| T2D PRS type | Sample | N | OR per PRS SD (95% CI) | P |
| --- | --- | --- | --- | --- |
| European-only (Khera *et al.*) | SJLIFE EUR | 3102 | 2.54 (1.92-3.35) | $4.5 \times 10^{-11}$ |
|  | SJLIFE AFR | 574 | 0.97 (0.39-2.41) | 0.95 |
| | CCSS EUR, original | 3902 | 2.16 (1.63-2.88) | $1.3 \times 10^{-7}$ |
| | CCSS EUR, expansion | 2063 | 3.06 (1.60-5.83) | $6.8 \times 10^{-4}$ |
| Multi-ancestry (Mahajan *et al.*) | SJLIFE EUR | 3102 | 1.84 (1.59-2.13) | $1.1 \times 10^{-16}$ |
| | SJLIFE AFR | 574 | 1.80 (1.22-2.64) | $2.8 \times 10^{-3}$ |
| | CCSS EUR, original | 3902 | 1.57 (1.36-1.81) | $7.8 \times 10^{-10}$ |
| | CCSS EUR, expansion | 2063 | 1.74 (1.25-2.42) | $9.5 \times 10^{-4}$ |

Abbreviations: SJLIFE, St. Jude Lifetime Cohort; CCSS, Childhood Cancer Survivor Study; EUR, European genetic ancestry; AFR, African genetic ancestry; PRS, polygenic risk score; RT, radiation therapy; OR, odds ratio; SD, standard deviation; CI, confidence interval; P, p-value.

The 338-variant multi-ancestry T2D PRS^11^ was further evaluated for potential treatment-related DM risk effect modification (Figure 2, Supplementary Table 10). No consistent evidence of variation in the odds of DM by abdominal RT exposures was observed (top vs. lowest PRS quintile in SJLIFE and CCSS EUR survivors with no abdominal RT exposures: OR_Q5_=4.90, P=3.6×10^-13^, versus with abdominal RT exposures ≥20 Gy: OR_Q5_=3.56, P=2.5×10^-5^). However, the multi-ancestry T2D PRS was consistently associated with increasing odds of DM among EUR and AFR survivors treated with alkylating agents (Figure 2). The top T2D PRS quintile was associated with 4.1-fold greater odds of DM (95% CI: 2.16-7.69; P=1.5×10^-5^) in SJLIFE and CCSS EUR survivors who were not treated with alkylating agents, whereas an 8.4-fold greater odds of DM (95% CI: 4.06-17.53; P=1.1×10^-8^) was observed in those with the same level of polygenic risk but were treated with alkylating agent doses ≥4,000 mg/m^2^. A similar trend was seen in SJLIFE AFR survivors, where those in the top T2D PRS quintile who were treated with any alkylating agents had 13.9-fold greater DM odds (95% CI: 1.24-154.48, P=0.033) than those in the lowest.

### Rare pathogenic variant burden in the *XNDC1N* gene and DM risk

For genome-wide gene-based rare variant burden analyses using pathogenic/likely pathogenic (P/LP) variants, we randomly split the SJLIFE EUR cohort *a priori* into discovery (N=1,550) and replication (N=1,552) datasets (see Methods). In the rare variant association analysis, we identified two genes with P/LP variant burden enrichment suggestively associated with DM in the SJLIFE EUR discovery sample (N=1,550; P<1.0×10^-3^): *XNDC1N* (XRCC1 N-terminal domain containing 1, N-terminal like; burden test P=8.5×10^-4^) and *ZNF655* (zinc finger protein 655; burden test P=7.1×10^-4^) (Supplementary Table 11). In the separate replication analysis (N=1,552), only *XNDC1N* remained significantly associated (P=7.1×10^-4^). In the combined data, the association between DM risk and *XNDC1N* P/LP burden was genome-wide significant (Bonferroni-corrected P<0.05/679 genes=7.4×10^-5^). Carrying either of the two *XNDC1N* P/LP variants conferred 8.65 greater odds of DM risk (MAF=0.003, 95% CI: 3.02- 24.74; burden test P=8.1×10^-6^) (Table 4). Single variant analyses further revealed that both *XNDC1N* P/LP variants contribute to increased DM risk (Supplementary Table 12). For secondary validation, we compared *XNDC1N* rare pathogenic risk allele frequencies among healthy EUR adults in the Genome Aggregation Database (gnomAD) to SJLIFE EUR survivor controls and cases (Supplementary Table 13). Compared to healthy gnomAD controls, the odds of carrying *XNDC1N* risk alleles among SJLIFE EUR controls were similar. In contrast, the odds of carrying these risk alleles were significantly higher among survivors with DM compared to healthy gnomAD controls (rs200142185: OR=4.72, 95% CI: 1.33-13.94, P=8.8×10^-3^; rs771623613, OR=19.77, 95% CI: 2.26-236.43, P=3.2×10^-3^).

**Table 4:** *XNDC1N* pathogenic/likely pathogenic rare variation enrichment association with diabetes mellitus risk among childhood cancer survivors in SJLIFE

| Rare variant association test | Discovery, SJLIFE EUR (N=1550) |  |  | Replication, SJLIFE EUR (N=1552) |  |  | Combined, SJLIFE EUR (N=3102) |  |  |
| --- | --- | --- | --- | --- | --- | --- | --- | --- | --- |
|  | MAF<br>(MAC) | OR<br>(95% CI) | P | MAF<br>(MAC) | OR<br>(95% CI) | P | MAF<br>(MAC) | OR<br>(95% CI) | P |
| Burden test, *XNDC1N* | 0.004<br>(13) | - | 8.5x10<sup>-4</sup> | 0.003<br>(8) | - | 9.4x10<sup>-3</sup> | 0.003<br>(21) | - | 8.1x10<sup>-6</sup> |
| Collapsed: ≥1 pathogenic variant in *XNDC1N* | 0.004<br>(13) | 8.80<br>(2.41-32.11) | 9.9x10<sup>-4</sup> | 0.003<br>(8) | 12.50<br>(1.99-78.65) | 7.1x10<sup>-3</sup> | 0.003<br>(21) | 8.65<br>(3.02-24.74) | 5.7x10<sup>-5</sup> |

Abbreviations: EUR, European genetic ancestry; MAF(C), minor allele frequency (count) across pathogenic/likely pathogenic variants mapped to gene; OR, odds ratio; CI, confidence interval; P, p-value.

### *In silico* functional annotation of DM risk variants

Functional annotations for all novel DM risk variants are provided in Supplementary Tables 14 and 15. The DM risk allele at the 8q11.21 locus index variant (rs6993752) is associated with *SNTG1*-region hypermethylation in BIOS QTL^19^ and overlaps putative promoter and enhancer chromatin states in liver and pancreas epigenomes. Index variants at 2p25.3, 5p15.2, and 19p12, all with evidence of trans-ancestral DM risk effects, overlap regions annotated with a Polycomb-repressed chromatin state in pancreatic cells. The 5p15.2 index variant risk allele is associated with increased alternative *FAM173B* splicing^20^, while the 19p12 index variant risk allele is associated with decreased transcription of nearby zinc finger proteins (*ZNF257*, *ZNF492*) in multiple tissues^20^. The *XNDC1N* rare pathogenic alleles are protein-truncating variants, overlapping transcribed regions in fat, liver, and pancreas epigenomes.

## DISCUSSION

Using a well-characterized cohort of long-term survivors of childhood cancer with clinically-ascertained DM status and WGS data, we identified five novel DM risk loci. Among these, we found that common risk variants at three loci, 5p15.2 (*LINC02112*), 2p25.3 (*MYT1L*), 19p12 (*ZNF492*), contributed to increased risk of DM in both EUR and AFR survivors treated with alkylating agents. In particular, AFR survivors with these DM risk alleles who were treated with alkylating agents experienced disproportionately greater odds of DM compared to their EUR counterpart. We also confirmed a general population 338-variant T2D PRS developed from a multi-ancestry genetic meta-analysis^11^ was potentially useful to survivors of AFR ancestry. Interestingly, this multi-ancestry T2D PRS was also associated with strikingly consistent increases in DM odds among EUR and AFR survivors treated with alkylating agents, contributing to its potential clinical utility for DM risk assessment in both ancestry groups. This study provides a step forward towards informing precision diabetes surveillance and survivorship care for all childhood cancer survivors, including those in minority ancestry groups.

Genetic and epigenetic regulation plausibly contribute to both inter-and intra-individual variation in cellular responses to alkylating agents^21, 22^, suggesting DM susceptibility can be influenced by both variations in genetic risk and exposures to alkylating agents. Notably, all three trans-ancestral DM risk loci identified in this study overlap putative Polycomb-repressed chromatin states in pancreatic cells. Polycomb group proteins have been shown to be involved in the epigenetic regulation of pancreatic β-cell replication, where the loss of Polycomb complex binding in pancreatic β-cells has been linked to reduced replication activity^23^. While it is unclear how the risk alleles identified in this study explicitly contribute to DM pathophysiology, these loci are strong candidates for further investigation. For example, rare deletions on the short arm of chromosome 2 (2p25.3) spanning the *MYT1L* gene have previously been linked to early-onset obesity and intellectual disability^24, 25^ and DM^26^. The 5p15.2 index variant is mapped to an intronic region of *LINC02112*. *LINC02112* includes variants suggestively associated with cytokine concentrations (interleukin-2 receptor antagonists), which have been linked to inflammatory diseases including DM^27^. Based on GTEx^20^ data, the 19p12 locus index variant is significantly associated with decreased expression of *ZNF257*. *ZNF257* variants common in East Asian populations but which are rare/monomorphic in European populations have been linked to T2D risk in a previous GWAS of 433,540 individuals of East Asian genetic ancestry^28^.

Interestingly, we also observed increased risk for DM associated with common variants at the 8q11.21 locus among overweight/obese EUR childhood cancer survivors. The index variant (rs6993752) did not appear to modify abdominal RT-or alkylating agent-related DM risks. This finding suggests DM-associated variants at 8q11.21 could increase risk though adiposity-related pathways, involving any of a myriad of risk factors for obesity, including exposures to cranial RT and glucocorticoids. The 8q11.21 index variant is located in an intronic region in *SNTG1* and is a methylation quantitative trait loci for this gene in whole blood in BIOS QTL^19^. While *SNTG1* has not been associated with DM risk in previous GWAS, other studies have identified *SNTG1* as a blood pressure locus^29^, a phenotype strongly linked to obesity.

We conducted the first genome-wide rare variant burden association analysis for DM in survivors, leading to the identification of *XNDC1N* (XRCC1 N-terminal domain containing 1) as a novel potential DM risk locus. These results were further supported by comparisons of *XNDC1N* risk allele frequencies between survivor controls and cases and healthy general population (gnomAD) adults. *XNDC1N* is predicted to encode XRCC1 N-terminal domain-like proteins, which have been shown contribute to XRCC1 protein function and the DNA repair process^30^. Specifically, XRCC1 N-terminal domain proteins modulate XRCC1 activity and are directly involved in the repair of single-strand DNA breaks, preferentially binding to single-strand break DNA^31, 32^. There is evidence that mutations in the *XRCC1* gene in cell lines show characteristics of DNA repair defects consistent with exposures to ionizing radiation, methylating reagents, and alkylating agents^33, 34^. While mutations in *XNDC1N* have not been linked to radiation or chemotherapy exposures in human populations, polymorphisms in *XRCC1* have been associated with radiation-induced late toxicity among breast cancer^35^ and prostate cancer patients^36^. Given the *XNDC1N* locus is putatively transcribed in fat, liver, and pancreas epigenomes, protein-truncating variants in *XNDC1N* may plausibly contribute to DM susceptibility among survivors.

The main strengths of our analysis include clinical ascertainment of DM status, rather than self-report, among SJLIFE participants, inclusion of a non-European genetic ancestry survivor population in analyses, and genome-wide interrogation of rare P/LP burden and DM risk. Despite these strengths, our analysis had several limitations. First, both our common and rare variant analyses are limited by the lack of secondary replication in independent cohorts of survivors with clinically ascertained DM status and WGS-based genotype data with sufficient sample size. However, such datasets do not currently exist. To minimize the possibility of false positive findings, we implemented study designs with separate replication studies, i.e., replication between ancestry subgroups in SJLIFE and secondary replication in CCSS for common variant GWAS, and an *a priori* discovery-replication data split for rare variant analysis. Second, our analyses have limited statistical power to formally test for gene-treatment statistical interactions. Top associations from variant main effects analyses may not overlap with top findings from a genome-wide analysis of gene-treatment interactions. However, the plausibility of treatment-and adiposity-related DM risk effects was supported by replicating stratified results across multiple study samples. Finally, *in silico* functional annotation is not a substitute for experimental functional validation. Thus, biological follow-up of these findings to elucidate their contributions to DM pathophysiology among childhood cancer survivors is warranted.

In summary, we identified five novel risk loci for DM, including a novel gene enriched with rare DM-associated protein-truncating variants, among childhood cancer survivors. The DM susceptibility conferred by these loci appeared to be modulated in part by survivors’ cancer treatment exposures. Trans-ancestry genetic analyses revealed three of these novel DM loci were associated with disproportionately greater alkylating agent-related risk among African-ancestry survivors. We also observed that a general population T2D PRS based on analyses in European samples did not appear to be informative for assessing DM risks in African-ancestry survivors, but a general population multi-ancestry T2D PRS may be relevant to more diverse populations of survivors, which may in part be due to its potential modification of alkylating agent-related DM risks across ancestral groups. Further investigations are needed to understand how these genetic risk factors contribute to DM risk mechanisms and can be utilized to support DM risk surveillance among survivors of diverse ancestry.

## METHODS

### Study population

For this study, we used data from childhood cancer survivors enrolled in two large retrospectively constructed cohort studies with prospective follow-up of late outcomes: the St. Jude Lifetime Cohort^37^ (SJLIFE), for discovery analyses, trans-ancestral replication, and trans-ancestry meta-analysis, and the Childhood Cancer Survivor Study^38^ (CCSS), for EUR-specific GWAS replication and secondary replication of findings from trans-ancestry meta-analysis. This study was approved by the institutional review boards at all participating study centers, including St. Jude Children’s Research Hospital (SJCRH). All participants provided written informed consent.

#### St. Jude Lifetime Cohort

Details of the SJLIFE study methodology have been described previously^37^. Briefly, SJLIFE includes five-year childhood cancer survivors who were treated for pediatric cancer at SJCRH between 1962 and 2012. All participants included in this investigation underwent at least one on-campus comprehensive medical evaluation and provided a biospecimen for DNA sequencing.

#### Childhood Cancer Survivor Study

Study design and methods for CCSS have been published in detail elsewhere^39, 40^. Participants include five-year childhood cancer survivors diagnosed at <21 years of age between 1970 and 1986 (described as the CCSS Original Cohort) and between 1987 and 1999 (described as the CCSS Expansion Cohort) who received treatment for pediatric cancer at one of 31 participating study institutions in the United States and Canada. In this analysis, we included all participants who completed a CCSS questionnaire which asked about the presence of chronic health conditions and provided a biospecimen for DNA genotyping or sequencing.

### Clinical risk factors and DM ascertainment

Data related to cancer treatments were abstracted from the medical record. Alkylating agent cumulative dose within five years of primary cancer diagnosis was abstracted and quantified as cyclophosphamide-equivalent dose^41^. Using data from the review of individual RT records, abdominal RT dose was estimated from the maximum tumor dose or total delivered dose from all overlapping abdominal RT fields^42^. Body mass index (BMI) for SJLIFE participants was calculated from height and weight measurements taken during the most recent campus study visit and adjusted for amputation, while BMI for CCSS participants was calculated from self-reported height and weight measurements from the most recently completed survey.

In SJLIFE, diabetes mellitus (DM) status was clinically ascertained. Participants with symptomatic diabetes, use of oral medications or insulin to treat diabetes, or blood tests consistent with DM diagnosis (fasting blood glucose ≥126 mg/dL, random glucose ≥200 mg/dL or HbA1c ≥6.5%) were considered to be DM cases in accordance with the NCI Common Terminology Criteria for Adverse Events (CTCAE) v4.03 classification system^43^ (CTCAE grades ≥2). While the prevalence of type 1 diabetes is low in survivors^5, 6^, further steps were taken to reduce the probability of including DM cases that were unlikely to be T2D. Clinical information was used to exclude participants with transient treatment-related hyperglycemia requiring intervention, as well as participants reporting insulin use and a DM diagnosis before or within 5 years of childhood cancer diagnosis.

In CCSS, we used case definitions of DM consistent with the literature^5, 6^ based on self-reported responses from all available baseline and follow-up questionnaires. Survivors who self-reported ever using an oral diabetes medication or insulin to treat diabetes were considered to be cases. Given the potential for misclassification, we also evaluated cases defined by self-report of ever using insulin to treat diabetes in sensitivity analyses.

### Genotype data

Methods used to generate genotype data in SJLIFE and CCSS have been described extensively elsewhere^44–48^. In brief, paired-end WGS was performed using Illumina HiSeq X10 or NovaSeq sequencers for all participants in SJLIFE and the CCSS Expansion Cohort, generating data for common/rare variant association analyses. For the CCSS Original Cohort, DNA was genotyped using the Illumina HumanOmni5Exome array and imputed with the Haplotype Reference Consortium r1.1 reference panel using Minimac3^49^; these data were used for analyses of common variants only. Sample and variant quality control measures were performed separately for the CCSS and SJLIFE autosomal variant data (Supplemental Methods). For each cohort, we identified participants of either European (EUR) or African (AFR) genetic ancestry using EIGENSTRAT-based principal components analyses of independent common variants implemented in PLINK^50^ version 1.9 (Supplemental Methods).

### Discovery of common trans-ancestral DM risk loci in SJLIFE

We first conducted ancestry-specific discovery GWAS for clinically-ascertained DM risk in SJLIFE, separately analyzing 3,102 EUR and 574 AFR survivors with complete phenotype and clinical covariate data. In each ancestral sample, common variants were evaluated (minor allele frequency or MAF ≥1%), including up to 10.5 million variants after excluding variants with excess missingness (>5%) and departures from Hardy-Weinberg equilibrium (P<1×10^-6^ among controls). Similar to previous GWAS^15^, logistic regression models of DM prevalence adjusted for sex, age at last contact, BMI, age at primary cancer diagnosis, cumulative doses of alkylating agents and abdominal RT, and the first ten ancestry-specific principal components were evaluated, assuming an additive genetic effect model.

To facilitate discovery of common loci with multi-ancestral DM risk effects in survivors, we conducted trans-ancestral replication in SJLIFE: top risk associations identified in AFR survivors were subsequently evaluated in EUR survivors and vice versa, using the same models applied in the discovery ancestry-specific GWAS. We prioritized variants from ancestry-specific discovery analyses with association test p-values <5×10^-6^ that were common (MAF ≥1%) across ancestral samples within a 100-kb window of the index variant (i.e., smallest p-value). Common variants meeting a p-value threshold accounting for Bonferroni-corrected multiple testing (0.05/number of tested variants in the locus with the index variant identified in discovery analysis) were considered to be replicated. In addition, trans-ancestry meta-analysis using the fixed-effects inverse variance-weighted method implemented in METAL^51^ was performed for index variants. Heterogeneity across genetic ancestral groups was examined using Cochran’s Q test^52^ and I^2^ index^53^. Meta-analysis for variants with evidence of ancestral allelic heterogeneity (P_het_ <0.05) was performed using the Han-Eskin random-effects model (RE2) in METASOFT^54^.

### Replication of common DM risk loci in CCSS

Replication was conducted in CCSS EUR survivors (excluding participants also enrolled in SJLIFE) for loci achieving genome-wide significance (P<5×10^-8^) in EUR-specific GWAS or the trans-ancestry meta-analysis, using the same models applied in SJLIFE. Separate replication analyses were conducted in the CCSS Original (N=3,902) and Expansion (N=2,063) cohorts due to differences in the derivation of genotype data. If the index variant was unavailable for evaluation in CCSS, we considered the next best variant proxy with the highest estimate of linkage disequilibrium (LD) with the index variant in the 1000 Genomes reference population. Replication was defined by association test p-values <0.05 in either the CCSS Original or Expansion cohorts.

### Treatment-and adiposity-stratified analyses of common DM risk loci

Similar to previous GWAS of late effects in survivors^55^, we evaluated treatment-or adiposity-stratified genetic associations with DM risk exclusively among genome-wide significant index variants with replication evidence to identify whether selected risk factors potentially modified risks for DM in survivors. We adopted this strategy to control the type I error probability while maximizing power for discovery in small survivor cohorts instead of performing genome-wide scans of gene-treatment interactions, which suffer losses in statistical power with the burden of multiple testing especially when considering several cancer treatment classes and dose definitions. Therefore, we examined replicated index variant associations with DM stratified by the two types of cancer treatments linked to DM^5, 6^: exposures to abdominal RT (none versus any) and alkylating agents (none versus any and lower versus higher dose [≥4,000 mg/m^2^] among those with no abdominal RT exposures). Due to the limited sample size of SJLIFE AFR survivors, alkylating agent-stratified analyses were not further stratified by abdominal RT exposures in this sample. Notably, obesity is strongly associated with treatment exposures and DM risk^6^, but abdominal RT is correlated with lower adiposity among survivors^17^. Because some DM-associated loci in survivors may not modify risks linked to abdominal RT or alkylating agents but instead modify obesity-associated risks for DM (e.g., involving cranial irradiation), we also conducted adiposity-stratified analyses (BMI≤25 kg/m^2^ versus being overweight or obese, BMI>25 kg/m^2^) for loci showing no clear DM risk effect heterogeneity with exposures to these two treatments. All statistical models assessed were adjusted for the same covariates, only omitting specific covariates directly related to the stratification type as appropriate.

### External T2D polygenic risk scores (PRSs)

For each survivor, we computed two published T2D PRSs derived from GWAS performed in non-cancer populations. First, we calculated a ∼6.9 million variant “European ancestry-only” PRS, derived from a European-ancestry T2D GWAS with 26,676 cases and 132,532 controls^56^, which was subsequently developed and validated in the predominantly European UK Biobank cohort^18^ (PGS Catalog: PGS000014). We also calculated a “multi-ancestry” PRS with 338 variants from a GWAS meta-analysis with 180,834 cases and >1.1 million controls (∼49% non-European ancestry)^11^. We implemented recommended best practices for PRS computation^57^ to calculate all PRSs using original publication weights to support comparability across studies, including common biallelic risk variants (minor allele frequency ≥1%) of unambiguous strand. All PRSs were generated in our data using PLINK v1.90b (--score)^50^. Adjusted associations between DM risk and standard deviation increases in scaled PRS values and PRS quintiles were evaluated among SJLIFE and CCSS survivors.

### Genome-wide analysis of genes enriched with pathogenic rare variant burden

For genome-wide gene-based rare variant burden analyses, we randomly split the SJLIFE EUR cohort *a priori* into approximately equal-sized discovery (N=1,550) and replication (N=1,552) datasets using the median last contact follow-up date (October 15, 2019). Given our limited sample size for discovery, we conducted gene-based rare variant burden tests^58–61^, an approach that compares the number of individuals carrying specific types of rare variants at a given gene among cases and controls and which is most powerful when the set of rare variants affect risk in the same direction and with similar magnitude^62^, using EPACTS^63^. Specifically, we considered rare variants (MAF<1% and minor allele counts ≥3) that were annotated as pathogenic/likely pathogenic (P/LP) based on functional/deleteriousness prediction algorithms, aggregated by gene with respect to the Ensembl^64^ (release 105) gene model (see Supplementary Methods). Associations with DM risk for genes enriched with protein-altering variant burden (with ≥2 variants per gene) with discovery association test p-values <1.0×10^-3^ were prioritized and were considered to be replicated if the gene-based P/LP burden met a p-value threshold accounting for multiple comparisons (0.05/number of prioritized genes from the discovery analysis) in the replication set. Among genes with replicated rare variant burden test associations, the p-value threshold for genome-wide significance for the combined study sample was set at P<7.4×10^-5^, accounting for a total of 679 genes with protein-altering variant burden enrichments.

### Bioinformatics analyses of genome-wide significant DM risk loci

We examined genome-wide significant index variants that were successfully replicated for associations with diabetes mellitus risk in the NHGRI-EBI GWAS Catalog^65^. Index variant associations in the general population using publicly available GWAS summary statistics for EUR participants in the UK Biobank Study^66^ for both self-reported T2D (N=361,141, with 2,292 cases) and ICD-10-coded T2D (N=361,194, with 888 cases) were extracted (http://www.nealelab.is/uk-biobank/). *In silico* functional/regulatory annotations for all genome-wide significant DM risk loci were obtained using external genomic data resources (Supplemental Methods).

### Data availability

Genotype and phenotype data for all SJLIFE survivors and a subset of CCSS survivors whose childhood cancer was diagnosed between 1987 and 1999 are accessible through the St. Jude Cloud (https://stjude.cloud). For CCSS survivors diagnosed between 1970 and 1986, genotype data are available through the database of Genotypes and Phenotypes (dbGaP accession number: phs001327.v2.p1) and relevant phenotype data may be requested through https://ccss.stjude.org/.

## Acknowledgements

This study was supported by the US National Cancer Institute (R21 CA261833, C Im/Y Yuan principal investigators; R01 CA216354, Y Yasui/J Zhang principal investigators; U01 CA195547, MM Hudson/K Ness, principal investigators; R01 CA261898, P Burridge/Y Sapkota, principal investigators; U24 CA55727, GT Armstrong, principal investigator; Cancer Center Support [CORE] Grant CA21765, C Roberts, principal investigator), and the American Lebanese Syrian Associated Charities.

