## Supplemental Materials for "Trans-ancestral genetic study of diabetes mellitus risk in survivors of childhood cancer: a report from the St. Jude Lifetime Cohort and the Childhood Cancer Survivor Study"

### SUPPLEMENTARY METHODS

*Genotype data quality control procedures in CCSS and SJLIFE*

For SJLIFE participants and participants in CCSS whose primary cancer was diagnosed between 1987 and 1999 (Expansion Cohort), paired-end whole-genome sequencing was performed using the Illumina HiSeq X10 or NovaSeq sequencers. Sequencing reads were mapped to the GRCh38 human genome assembly using the Burrows-Wheeler Aligner algorithm[1] (BWA-MEM). Variant calls were processed with GATK v3.4.0[2] and BCFtools[3] using recommended best practices. PLINK v1.90b[4] and VCFtools v0.1.13[5] were used to perform additional quality control, applying the following sample exclusion criteria: excess missingness (≥5%), cryptic relatedness (pi-hat>0.25), and excess heterozygosity (>3 standard deviations from sample mean). Variants with minimum genotype quality score <20, minimum mean read depth <10, HWE test $P<1\times10^{-10}$ and >10% missingness across samples were removed.

For CCSS participants whose primary cancer was diagnosed before 1987 (Original Cohort), DNA was genotyped at the Cancer Genomics Research Laboratory of the National Cancer Institute (Bethesda, MD) using the Illumina HumanOmni5Exome array. Genotypes were called with Genotyping Module v1.9 (Illumina GenomeStudio software v2011.1). Samples with excess missingness (≥8%), heterozygosity (<0.11 or >0.16), sex discordance (X chromosome heterozygosity >5% for males or <20% for females), and cryptic relatedness (identity-by-descent sharing for first relatives, i.e., >0.70) were removed. Variants with Hardy Weinberg equilibrium (HWE) test P<0.001, call rates ≤97%, and of ambiguous strand (A/T or C/G) were removed. After this quality control process, genotypes were imputed via the Michigan Imputation Server using Minimac3[6] and the Haplotype Reference Consortium r1.1 reference panel. Genetic variants with low imputation quality ($r^2$<0.8) were removed.

*Genetic ancestry*

Procedures to identify the genetic ancestry of SJLIFE and CCSS samples have been described elsewhere.[7,8] Briefly, an EIGENSTRAT-based principal component analysis[9] was performed with PLINK v1.90b for each cohort by combining cohort samples with samples from 1000 Genomes (1000G) global reference populations. Samples with principal component scores ±3 SD of the means of the first two principal components in the 1000G European or African populations were considered to be of European or African ancestry, respectively. Principal components were then computed using independent common variants for each genetically inferred ancestral population in SJLIFE and CCSS for inclusion in multivariable models.

*Variant annotation for rare variant association analyses*

Methods to pool or collapse rare variants are instrumental in association studies of rare variation, enhancing power by examining the combined effect of different rare variants at a given locus[10-12]. To this end, genic annotations were obtained from the ANNOVAR[13] based on the RefSeq gene model, which included the 3' untranslated region (UTR) and 5' UTR, and intron, splicing, frameshift, synonymous, non-synonymous, stopgain, stoploss and upstream (within 5 kilobases of the start of a gene) regions. We categorized rare/low-frequency variants as pathogenic or likely pathogenic that were: (1) protein truncating variants (PTVs), or coding variants that are annotated to cause frameshift, stoploss, stopgain and splicing site changes as predicted by by SnpEff[14] (version 5.1d); or (2) pathogenic or likely-pathogenic per ClinVar[15] predictions.

*Functional/regulatory annotations of genetic variants*

Coding and regulatory consequences of replicated index variants, including Combined Annotation Dependent Depletion[16] (CADD) scores predicting variant deleteriousness, were annotated using the Ensembl Variant Effect Predictor[17] (VEP v99). Significant associations with gene expression, i.e., expression quantitative trait loci (eQTLs; FDR<5%), and DNA methylation levels, i.e., methylation quantitative trait loci (meQTLs; FDR<5%), were identified from the Genotype-Tissue Expression[18] (GTEx v8) project, eQTLGen Consortium[19], and BIOS Consortium[20] (BIOS QTL) databases. Chromatin state annotations for regulatory states (e.g., promoters, enhancers, transcription, Polycomb repressed) based on the 15-state ChromHMM model trained on 12 epigenetic marks for 127 epigenomes[21] were obtained from the Roadmap Epigenomics Consortium. We used pooled chromatin state annotations for any promoter (states 1-2, 10), transcription (states 3-5), enhancer (states 6-7, 11-12), and polycomb repressed (states 13-14) states among relevant epigenomes (liver [E066, E118]; pancreas [E087, E098]; fat [E023, E025, E063]).

### SUPPLEMENTARY FIGURES AND TABLES

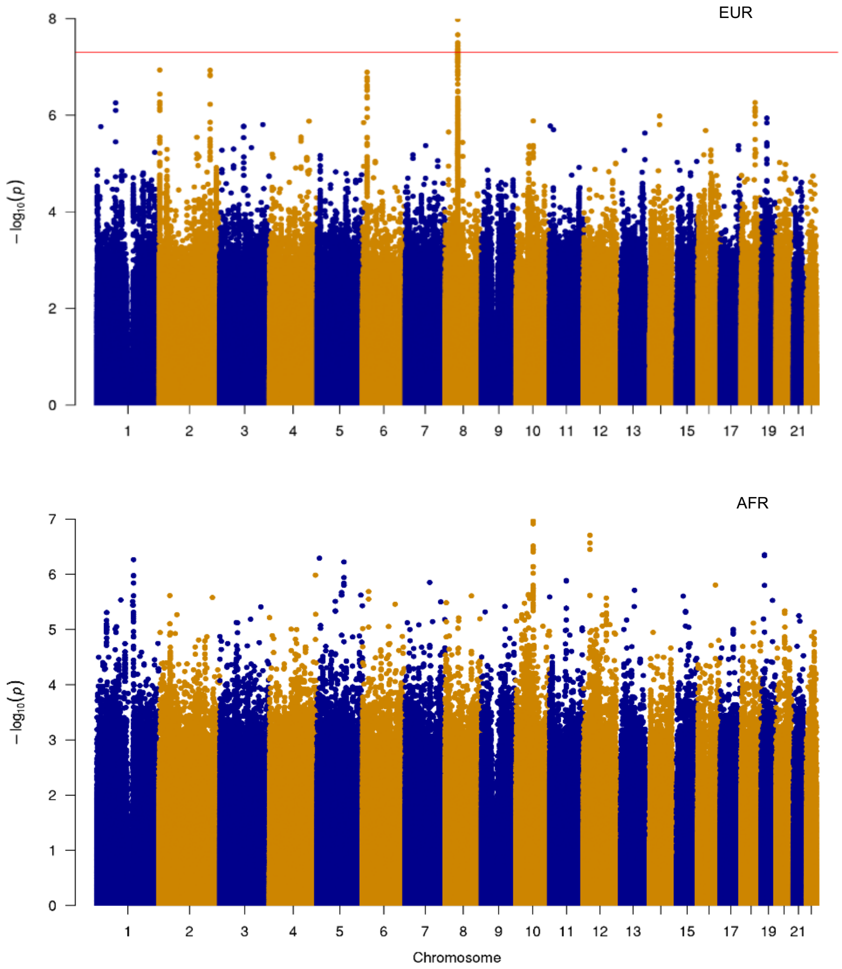


**Supplementary Figure 1**: Manhattan plots for SNP association test p-values among SJLIFE survivors of European (EUR; top panel) and African (AFR; bottom panel) genetic ancestry. The Manhattan plots illustrate $-\log_{10}$ p-values for SNP associations with diabetes mellitus risk (y-axis) against SNP genomic positions (x-axis). The solid red line signifies the genome-wide significance threshold (log-transformed $P=5 \times 10^{-8}$).

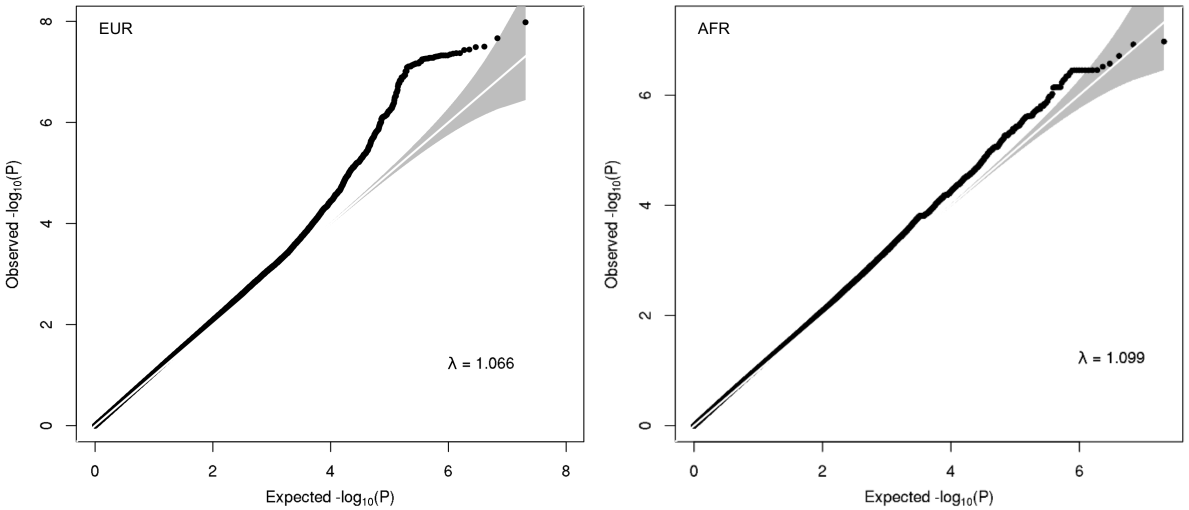


**Supplementary Figure 2**: Quantile-quantile (QQ) plot for SNP association test p-values from the SJLIFE discovery GWAS. European (EUR) and African (AFR) genetic ancestry QQ plots are shown on the left and right panels, respectively. The QQ plots show observed $-\log_{10}$ p-values (y-axis) against those expected under the null distribution of no association (x-axis), as well as the genomic inflation factor (lambda).

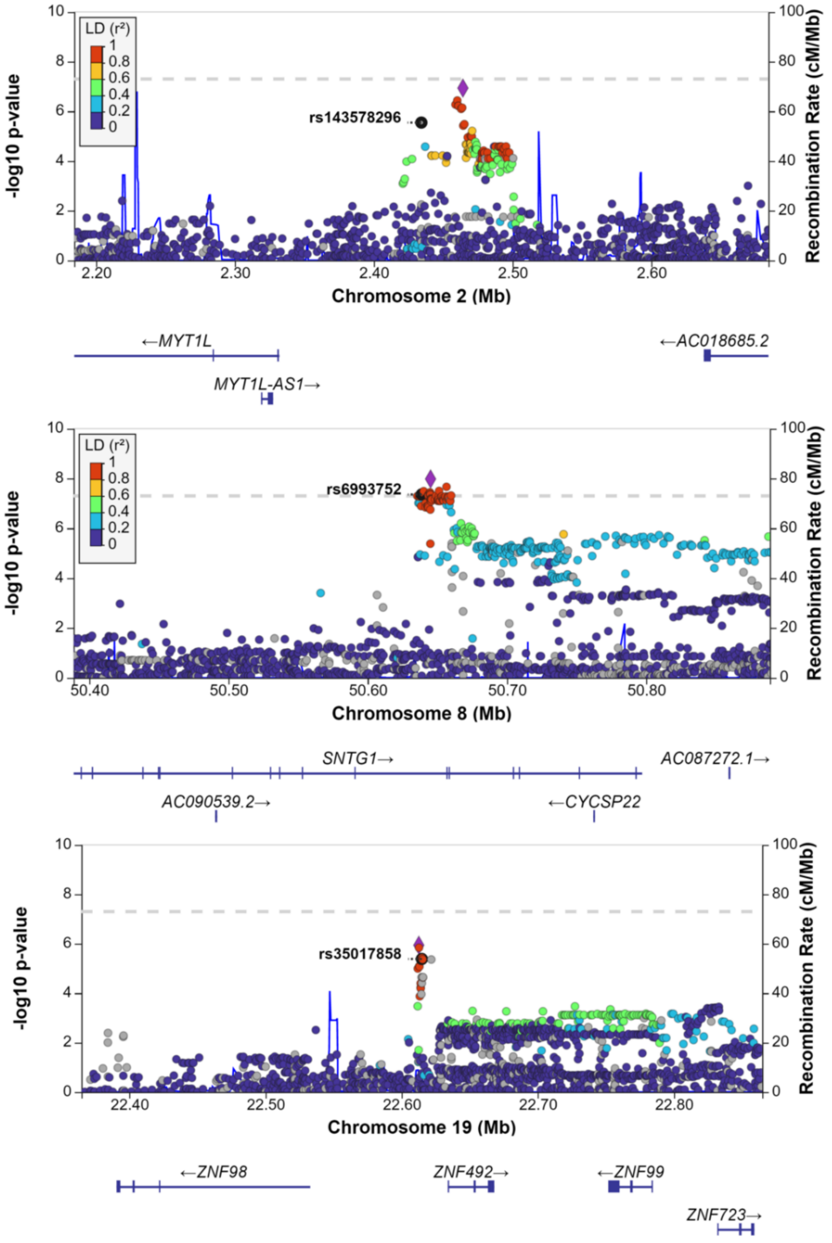


**Supplementary Figure 3:** LocusZoom plots of novel diabetes mellitus risk loci from common variant association analyses among SJLIFE survivors of European ancestry. Single variant association test p-values from discovery analyses are shown (-$\log_{10}$ scale) for variants within a 500-kilobase window around the index variant at each locus (annotated in each panel). SNPs are color coded by their magnitude of linkage disequilibrium (LD, measured as $r^2$) with the most significantly associated variant in the window in the 1000 Genomes European reference panel.

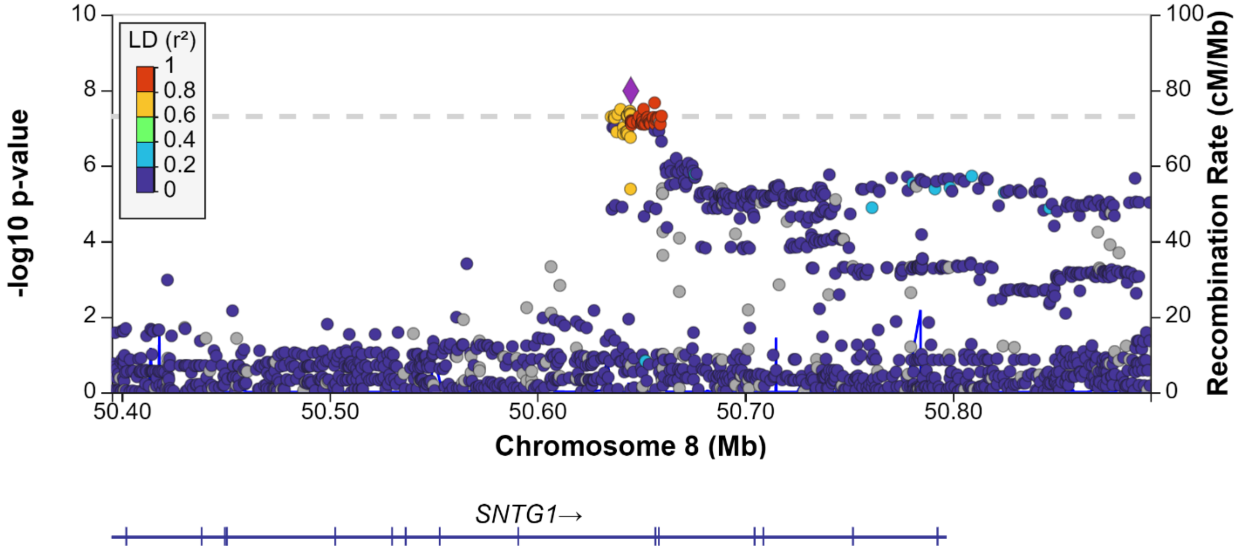


**Supplementary Figure 4**: LocusZoom plot of the novel diabetes mellitus risk loci at 8q11.21 with African ancestry linkage disequilibrium patterns. SNPs are color coded by their magnitude of linkage disequilibrium (LD, measured as $r^2$) with the most significantly associated variant in the window in the 1000 Genomes African reference panel.

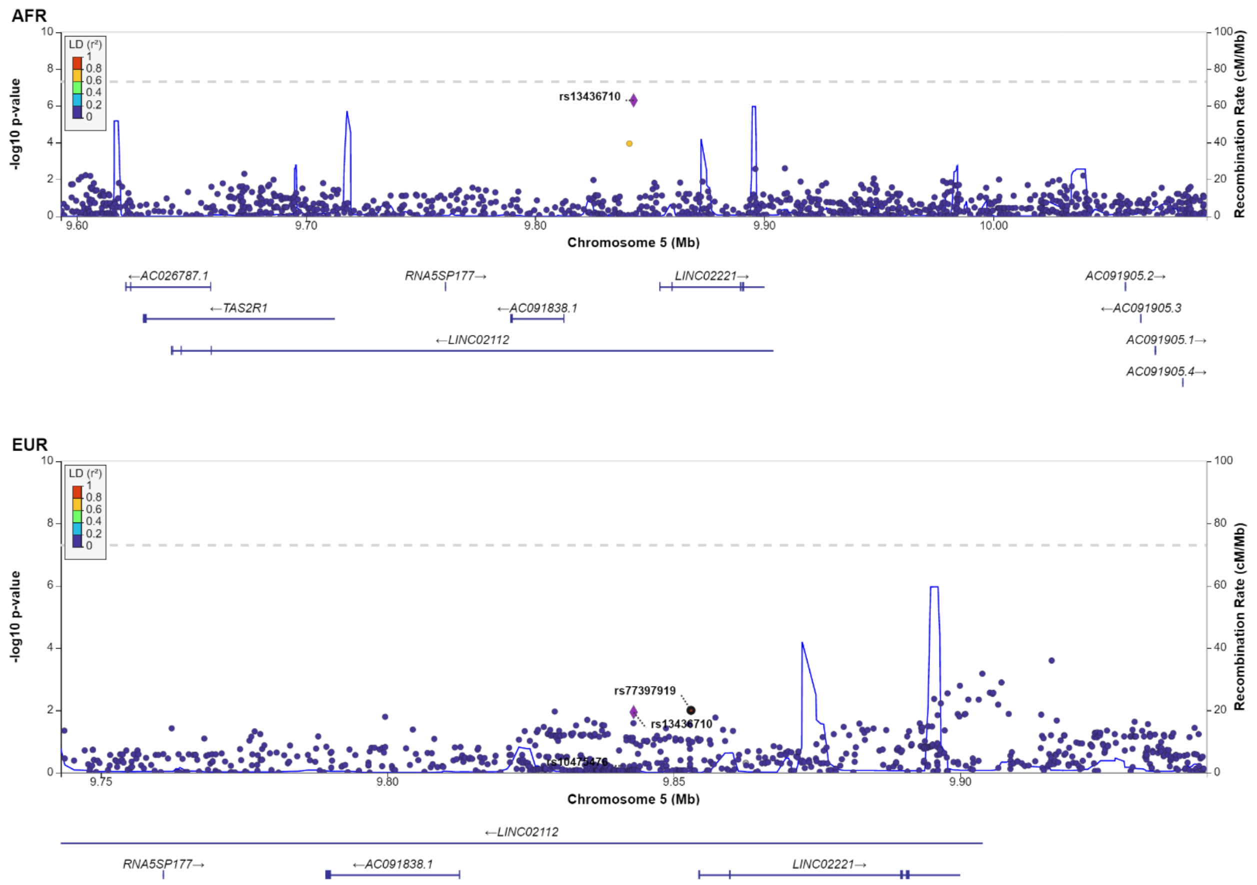


**Supplementary Figure 5**: LocusZoom plot of the novel diabetes mellitus risk loci at 5p15.2 with African and European ancestry linkage disequilibrium patterns. SNPs are color coded by their magnitude of linkage disequilibrium (LD, measured as $r^2$) with the most significantly associated variant in the window in the SJLIFE AFR discovery GWAS, in both SJLIFE AFR (top) and EUR (bottom) samples.

**Supplementary Table 1**: Multivariable model for diabetes mellitus risk with clinical risk factors only for SJLIFE participants of European or African genetic ancestry

| | SJLIFE EUR (N=3,102) | | | SJLIFE AFR (N=574) | | |
|---|---|---|---|---|---|---|
| Variables | OR | 95% CI | P | OR | 95% CI | P |
| Sex | 1.23 | 0.94-1.62 | 0.13 | 0.81 | 0.40-1.63 | 0.56 |
| Age at cancer diagnosis, per 5 years | 0.94 | 0.83-1.06 | 0.29 | 0.61 | 0.43-0.87 | $6.5 \times 10^{-3}$ |
| Age at follow-up, per 10 years | 1.91 | 1.67-2.19 | $1.2 \times 10^{-20}$ | 2.52 | 1.80-3.54 | $8.0 \times 10^{-8}$ |
| Adjusted body mass index, per 5 $kg/m^2$ | 1.56 | 1.44-1.69 | $3.8 \times 10^{-28}$ | 1.45 | 1.18-1.78 | $3.9 \times 10^{-4}$ |
| Maximum cumulative abdominal radiation dose, per 10 Gy | 1.23 | 1.10-1.38 | $3.6 \times 10^{-4}$ | 1.23 | 0.91-1.66 | 0.17 |
| Alkylating agent CED dose, per $g/m^2$ | 0.99 | 0.97-1.01 | 0.24 | 0.95 | 0.89-1.01 | 0.11 |
| Any alkylating agents[a] | 1.35 | 1.02-1.80 | 0.039 | 0.52 | 0.25-1.08 | 0.081 |

Abbreviations: SJLIFE, St. Jude Lifetime Cohort Study; EUR, European genetic ancestry; AFR, African genetic ancestry; m, meters; g, grams; kg, kilograms; Gy, Gray; CED, cyclophosphamide equivalent dose; OR, odds ratio; CI, confidence interval; P, p-value.

a. Adjusted ORs are based on multivariable models with the same set of adjustment covariates used in all genetic analyses (see previous rows), with the exception of a threshold-dose variable definition for alkylating agents (receipt of any alkylating agents vs. none).

**Supplementary Table 2**: Demographic and clinical characteristics of childhood cancer survivors in CCSS

| | CCSS original cohort (N=3902) | | CCSS expansion cohort (N=2063) | |
|---|---|---|---|---|
| Characteristic | N | % | N | % |
| Sex | | | | |
| Male | 1862 | 47.7 | 966 | 48.0 |
| Female | 2040 | 52.3 | 1097 | 52.0 |
| Age at follow up (in years) | | | | |
| <25 | 151 | 3.9 | 516 | 28.2 |
| 25-34 | 941 | 24.1 | 1100 | 54.3 |
| 35-44 | 1513 | 38.8 | 428 | 16.8 |
| ≥45 | 1297 | 33.2 | 19 | 0.7 |
| Body mass index (kg/m$^2$) | | | | |
| ≤25 | 1609 | 41.2 | 1002 | 48.8 |
| >25 to ≤30 | 1268 | 32.5 | 599 | 29.6 |
| >30 to ≤40 | 834 | 21.4 | 387 | 18.8 |
| >40 | 191 | 4.9 | 75 | 2.9 |
| Developed diabetes mellitus[a] | 223 | 5.7 | 47 | 2.3 |
| Age at cancer diagnosis (in years) | | | | |
| <5 | 1536 | 39.4 | 721 | 40.4 |
| 5-9 | 848 | 21.7 | 485 | 26.1 |
| 10-14 | 819 | 21.0 | 520 | 20.5 |
| ≥15 | 699 | 17.9 | 337 | 13.0 |
| Childhood cancer diagnosis | | | | |
| Leukemias | 1187 | 30.4 | 442 | 39.4 |
| Central nervous system tumors | 553 | 14.2 | 528 | 19.7 |
| Hodgkin lymphoma | 475 | 12.2 | 248 | 9.3 |
| Non-Hodgkin lymphoma | 306 | 7.8 | 191 | 7.1 |
| Kidney tumors | 405 | 10.4 | 203 | 7.6 |
| Soft tissue sarcomas | 362 | 9.3 | 107 | 4.0 |
| Other | 614 | 15.7 | 344 | 12.9 |
| Cancer treatments | | | | |
| Any abdominal radiation therapy (RT) | 983 | 25.2 | 347 | 13.7 |
| Any alkylating agents | 1736 | 44.5 | 1083 | 52.7 |
| Dosages: Median (IQR)[b] | | | | |
| Abdominal RT tumor dose (cGy) | 983 | 3000 (2000-3600) | 347 | 2300 (1200-3550) |
| Alkylating agents (CED, mg/m$^2$) | 1736 | 8665 (4371-13919) | 1083 | 7167 (3948-12187) |

Abbreviations: CCSS, Childhood Cancer Survivor Study; IQR, interquartile range; RT, radiation therapy; CED, cyclophosphamide equivalent dose; mg, milligrams; m, meters; kg, kilograms. Percentages reflect primary cancer diagnosis sampling weights.

a. Self-reported diabetes mellitus requiring medication was evaluated in CCSS.

b. Treatment dose distribution statistics are provided for childhood cancer survivors who received any of the specified treatments and had non-missing dosage information.

**Supplementary Table 3**: SJLIFE trans-ancestry meta-analysis results for top genetic associations with diabetes mellitus risk (100-kb window, $P<5\times10^{-6}$)

| | | | | SJLIFE EUR | | | SJLIFE AFR | | | SJLIFE EUR + AFR meta-analysis | | |
|---|---|---|---|---|---|---|---|---|---|---|---|---|
| Chr | BP | EA | NEA | EAF | OR (95% CI) | P | EAF | OR (95% CI) | P | OR | P | $P_{het}$ |
| 2 | 2434309 | A | AAAATAAAATAAAATAAAATAAAAAC | 0.10 | 1.99 (1.49-2.66) | $2.8\times10^{-6}$ | 0.17 | 2.30 (1.30-4.07) | $4.2\times10^{-3}$ | 2.05 (1.59-2.65) | $4.5\times10^{-8}$ | 0.66 |
| 2 | 2460867 | C | T | 0.13 | 1.96 (1.51-2.56) | $6.0\times10^{-7}$ | 0.03 | 4.14 (1.35-12.69) | 0.013 | 2.04 (1.58-2.64) | $5.6\times10^{-8}$ | 0.20 |
| 2 | 2462940 | A | G | 0.13 | 1.95 (1.49-2.54) | $8.0\times10^{-7}$ | 0.03 | 4.24 (1.38-13.06) | 0.012 | 2.03 (1.57-2.63) | $7.4\times10^{-8}$ | 0.19 |
| 8 | 50635641 | T | C | 0.15 | 1.98 (1.55-2.54) | $5.1\times10^{-8}$ | 0.38 | 0.54 (1.31-0.93) | 0.027 | 1.60 (1.28-2) | $4.5\times10^{-5}$ | $2.3\times10^{-5}$ |
| 8 | 50636999 | C | T | 0.15 | 1.98 (1.55-2.53) | $5.6\times10^{-8}$ | 0.38 | 0.54 (1.31-0.93) | 0.027 | 1.59 (1.27-1.99) | $5.2\times10^{-5}$ | $2.2\times10^{-5}$ |
| 8 | 50637285 | A | T | 0.15 | 1.99 (1.55-2.54) | $4.7\times10^{-8}$ | 0.37 | 0.57 (1.33-0.98) | 0.042 | 1.61 (1.28-2.01) | $3.5\times10^{-5}$ | $4.1\times10^{-5}$ |
| 8 | 50637444 | A | G | 0.15 | 1.98 (1.55-2.53) | $5.4\times10^{-8}$ | 0.38 | 0.54 (1.31-0.93) | 0.026 | 1.59 (1.27-1.99) | $5.2\times10^{-5}$ | $2.1\times10^{-5}$ |
| 8 | 50638825 | G | A | 0.15 | 1.99 (1.56-2.55) | $4.4\times10^{-8}$ | 0.38 | 0.53 (1.30-0.91) | 0.022 | 1.59 (1.27-2) | $4.8\times10^{-5}$ | $1.6\times10^{-5}$ |
| 8 | 50641552 | A | G | 0.15 | 1.94 (1.52-2.48) | $1.4\times10^{-7}$ | 0.37 | 0.55 (1.32-0.95) | 0.032 | 1.57 (1.25-1.97) | $8.8\times10^{-5}$ | $4.0\times10^{-5}$ |
| 8 | 50643118 | T | G | 0.15 | 1.94 (1.52-2.48) | $1.4\times10^{-7}$ | 0.37 | 0.55 (1.32-0.95) | 0.032 | 1.57 (1.25-1.97) | $8.7\times10^{-5}$ | $4.1\times10^{-5}$ |
| 8 | 50644980 | A | G | 0.18 | 1.73 (1.37-2.18) | $4.2\times10^{-6}$ | 0.40 | 0.58 (1.34-0.99) | 0.046 | 1.45 (1.17-1.8) | $6.3\times10^{-4}$ | $2.4\times10^{-4}$ |
| 8 | 50660409 | G | GAAAA | 0.21 | 1.68 (1.35-2.09) | $4.0\times10^{-6}$ | 0.47 | 0.44 (1.25-0.77) | $4.5\times10^{-3}$ | 1.41 (1.15-1.73) | $1.0\times10^{-3}$ | $1.7\times10^{-5}$ |
| 8 | 50661918 | G | C | 0.30 | 1.66 (1.35-2.04) | $1.2\times10^{-6}$ | 0.64 | 0.59 (1.35-1.00) | 0.048 | 1.45 (1.2-1.75) | $1.3\times10^{-4}$ | $3.1\times10^{-4}$ |
| 8 | 50666979 | A | G | 0.21 | 1.74 (1.40-2.16) | $6.4\times10^{-7}$ | 0.51 | 0.57 (1.33-0.97) | 0.037 | 1.48 (1.21-1.81) | $1.3\times10^{-4}$ | $1.5\times10^{-4}$ |
| 8 | 50667110 | A | G | 0.21 | 1.71 (1.37-2.12) | $1.5\times10^{-6}$ | 0.60 | 0.57 (1.33-0.97) | 0.039 | 1.46 (1.19-1.79) | $2.3\times10^{-4}$ | $1.9\times10^{-4}$ |
| 8 | 50667850 | G | T | 0.21 | 1.71 (1.37-2.12) | $1.5\times10^{-6}$ | 0.61 | 0.57 (1.33-0.96) | 0.035 | 1.45 (1.19-1.78) | $2.7\times10^{-4}$ | $1.5\times10^{-4}$ |
| 8 | 50671413 | C | T | 0.21 | 1.72 (1.38-2.13) | $1.2\times10^{-6}$ | 0.61 | 0.56 (1.33-0.94) | 0.029 | 1.45 (1.19-1.78) | $2.7\times10^{-4}$ | $1.0\times10^{-4}$ |
| 8 | 50671938 | A | G | 0.21 | 1.69 (1.36-2.10) | $3.0\times10^{-6}$ | 0.19 | 0.44 (1.20-0.97) | 0.043 | 1.53 (1.24-1.9) | $7.3\times10^{-5}$ | $1.4\times10^{-3}$ |
| 8 | 50672410 | T | A | 0.21 | 1.72 (1.38-2.14) | $1.0\times10^{-6}$ | 0.61 | 0.55 (1.32-0.92) | 0.024 | 1.46 (1.19-1.78) | $2.5\times10^{-4}$ | $8.0\times10^{-5}$ |
| 8 | 50672830 | C | T | 0.21 | 1.73 (1.39-2.15) | $8.5\times10^{-7}$ | 0.51 | 0.57 (1.33-0.97) | 0.037 | 1.48 (1.21-1.81) | $1.6\times10^{-4}$ | $1.6\times10^{-4}$ |
| 8 | 50673224 | G | A | 0.21 | 1.71 (1.37-2.12) | $1.5\times10^{-6}$ | 0.61 | 0.55 (1.32-0.92) | 0.024 | 1.45 (1.18-1.77) | $3.2\times10^{-4}$ | $9.0\times10^{-5}$ |
| 8 | 50674706 | G | A | 0.21 | 1.72 | $9.9\times10^{-7}$ | 0.61 | 0.55 | 0.024 | 1.46 | $2.5\times10^{-4}$ | $7.9\times10^{-5}$ |

| | | | | SJLIFE EUR | | | SJLIFE AFR | | | SJLIFE EUR + AFR meta-analysis | | |
|---|---|---|---|---|---|---|---|---|---|---|---|---|
| Chr | BP | EA | NEA | EAF | OR (95% CI) | P | EAF | OR (95% CI) | P | OR | P | $P_{het}$ |
| | | | | | (1.39-2.14) | | | (1.32-0.92) | | (1.19-1.78) | | |
| 8 | 50675283 | G | A | 0.21 | 1.71 (1.37-2.12) | $1.4 \times 10^{-6}$ | 0.61 | 0.55 (1.32-0.92) | 0.024 | 1.45 (1.18-1.77) | $3.1 \times 10^{-4}$ | $8.9 \times 10^{-5}$ |
| 8 | 50676103 | C | T | 0.21 | 1.71 (1.37-2.12) | $1.7 \times 10^{-6}$ | 0.19 | 0.44 (1.20-0.98) | 0.044 | 1.55 (1.26-1.91) | $4.4 \times 10^{-5}$ | $1.3 \times 10^{-3}$ |
| 8 | 50676751 | G | T | 0.21 | 1.71 (1.37-2.12) | $1.6 \times 10^{-6}$ | 0.19 | 0.44 (1.20-0.98) | 0.044 | 1.55 (1.26-1.91) | $4.3 \times 10^{-5}$ | $1.3 \times 10^{-3}$ |
| 8 | 50681567 | G | A | 0.20 | 1.68 (1.34-2.09) | $5.0 \times 10^{-6}$ | 0.61 | 0.57 (1.34-0.96) | 0.034 | 1.42 (1.16-1.74) | $7.4 \times 10^{-4}$ | $1.9 \times 10^{-4}$ |
| 19 | 22614854 | G | A | 0.05 | 2.26 (1.60-3.20) | $4.1 \times 10^{-6}$ | 0.22 | 2.99 (1.66-5.37) | $2.6 \times 10^{-4}$ | 2.43 (1.80-3.28) | $5.7 \times 10^{-9}$ | 0.43 |
| 19 | 22614973 | A | G | 0.05 | 2.28 (1.61-3.24) | $3.7 \times 10^{-6}$ | 0.16 | 2.23 (1.17-4.25) | 0.014 | 2.27 (1.67-3.09) | $1.7 \times 10^{-7}$ | 0.95 |
| 19 | 22615242 | T | C | 0.05 | 2.26 (1.59-3.19) | $4.4 \times 10^{-6}$ | 0.22 | 2.64 (1.51-4.62) | $6.6 \times 10^{-4}$ | 2.36 (1.76-3.17) | $1.2 \times 10^{-8}$ | 0.64 |
| 19 | 22615378 | A | G | 0.05 | 2.26 (1.60-3.20) | $4.3 \times 10^{-6}$ | 0.23 | 2.43 (1.41-4.20) | $1.4 \times 10^{-3}$ | 2.31 (1.72-3.09) | $2.3 \times 10^{-8}$ | 0.82 |
| 19 | 22615432 | C | A | 0.05 | 2.26 (1.60-3.20) | $4.3 \times 10^{-6}$ | 0.23 | 2.43 (1.41-4.19) | $1.5 \times 10^{-3}$ | 2.31 (1.72-3.09) | $2.3 \times 10^{-8}$ | 0.83 |

Abbreviations: SJLIFE, St. Jude Lifetime Cohort Study; Chr, chromosome; BP, base position, GRCh38 (hg38) build; EA, effect (risk) allele; NEA, non-effect (reference) allele; EUR, European genetic ancestry; AFR, African genetic ancestry; OR, odds ratio; CI, confidence interval; P, p-value.

**Supplementary Table 4**: CCSS replication results for top genetic associations with risk for self-reported diabetes mellitus requiring oral medications or insulin (European genetic ancestry)

| | | | | CCSS Original (N=223 cases) | | | CCSS Expansion (N=47 cases) | | |
|---|---|---|---|---|---|---|---|---|---|
| Chr | BP | EA | NEA | EAF | OR (95% CI) | P | EAF | OR (95% CI) | P |
| 2 | 2460867 | C | T | 0.14 | 1.20 (0.91-1.59) | 0.19 | 0.13 | 1.03 (0.52-2.07) | 0.92 |
| 2 | 2462940 | A | G | 0.14 | 1.21 (0.92-1.60) | 0.17 | 0.13 | 1.03 (0.52-2.07) | 0.93 |
| 5 | 9843018 | C | T | | | | 0.01 | 1.02 (0.13-7.77) | 0.98 |
| 5 | 9841183 | G | C | 0.07 | 1.48 (1.06-2.07) | 0.021 | 0.07 | 0.79 (0.31-2.03) | 0.63 |
| 5 | 9853032 | C | G | | | | 0.01 | 0.96 (0.13-7.36) | 0.97 |
| 8 | 50635641 | T | C | 0.16 | 1.39 (1.09-1.78) | $9.0 \times 10^{-3}$ | 0.15 | 0.95 (0.52-1.73) | 0.88 |
| 8 | 50636999 | C | T | 0.16 | 1.39 (1.08-1.77) | $9.4 \times 10^{-3}$ | 0.15 | 0.95 (0.52-1.74) | 0.88 |
| 8 | 50637285 | A | T | 0.16 | 1.39 (1.08-1.77) | $9.4 \times 10^{-3}$ | 0.15 | 0.95 (0.52-1.73) | 0.87 |
| 8 | 50637444 | A | G | 0.16 | 1.39 (1.08-1.77) | $9.4 \times 10^{-3}$ | 0.15 | 0.95 (0.52-1.74) | 0.88 |
| 8 | 50638825 | G | A | 0.16 | 1.39 (1.08-1.77) | $9.4 \times 10^{-3}$ | 0.15 | 0.95 (0.52-1.74) | 0.88 |
| 8 | 50641552 | A | G | 0.16 | 1.39 (1.08-1.77) | $9.4 \times 10^{-3}$ | 0.15 | 0.95 (0.52-1.74) | 0.88 |
| 8 | 50643118 | T | G | 0.16 | 1.39 (1.08-1.77) | $9.4 \times 10^{-3}$ | 0.15 | 0.95 (0.52-1.74) | 0.88 |
| 8 | 50644980 | A | G | 0.18 | 1.26 (0.98-1.60) | 0.07 | 0.18 | 1.00 (0.58-1.73) | 1.00 |
| 8 | 50661918 | G | C | 0.69 | 0.89 (0.72-1.09) | 0.26 | 0.70 | 0.79 (0.54-1.14) | 0.21 |
| 8 | 50666979 | A | G | 0.22 | 1.20 (0.96-1.50) | 0.11 | 0.22 | 1.56 (1.01-2.40) | 0.045 |
| 8 | 50667110 | A | G | 0.22 | 1.21 (0.97-1.51) | 0.10 | 0.22 | 1.56 (1.01-2.39) | 0.044 |
| 8 | 50667850 | G | T | 0.22 | 1.21 (0.97-1.51) | 0.10 | 0.22 | 1.33 (0.84-2.10) | 0.23 |
| 8 | 50672410 | T | A | 0.22 | 1.21 (0.97-1.51) | 0.10 | 0.22 | 1.32 (0.83-2.10) | 0.23 |
| 8 | 50672830 | C | T | 0.22 | 1.21 (0.96-1.51) | 0.10 | 0.22 | 1.33 (0.84-2.11) | 0.23 |
| 8 | 50673224 | G | A | 0.22 | 1.21 (0.97-1.51) | 0.10 | 0.22 | 1.32 (0.83-2.10) | 0.23 |
| 8 | 50674706 | G | A | 0.22 | 1.21 (0.97-1.51) | 0.10 | 0.22 | 1.32 (0.83-2.10) | 0.23 |
| 8 | 50675283 | G | A | 0.22 | 1.21 (0.97-1.51) | 0.10 | 0.22 | 1.43 (0.91-2.26) | 0.12 |
| 8 | 50676103 | C | T | 0.22 | 1.2 (0.96-1.49) | 0.12 | 0.21 | 1.34 (0.84-2.14) | 0.22 |
| 8 | 50676751 | G | T | 0.22 | 1.19 (0.95-1.49) | 0.13 | 0.21 | 1.45 (0.91-2.30) | 0.11 |
| 8 | 50681567 | G | A | 0.21 | 1.17 (0.93-1.47) | 0.18 | 0.20 | 1.54 (0.97-2.45) | 0.07 |
| 19 | 22614854 | G | A | 0.06 | 1.17 (0.77-1.77) | 0.46 | 0.06 | 0.76 (0.31-1.87) | 0.55 |
| 19 | 22614973 | A | G | 0.06 | 1.17 (0.77-1.78) | 0.46 | 0.06 | 0.77 (0.31-1.91) | 0.57 |
| 19 | 22615242 | T | C | 0.06 | 1.17 (0.77-1.77) | 0.46 | 0.06 | 0.76 (0.31-1.88) | 0.55 |
| 19 | 22615378 | A | G | 0.06 | 1.17 (0.77-1.77) | 0.46 | 0.06 | 0.76 (0.31-1.87) | 0.55 |
| 19 | 22615432 | C | A | 0.06 | 1.17 (0.77-1.77) | 0.46 | 0.06 | 0.76 (0.31-1.88) | 0.55 |

Abbreviations: CCSS, Childhood Cancer Survivor Study; Chr, chromosome; BP, base position, GRCh38 (hg38) build; EA, effect (risk) allele; NEA, non-effect (reference) allele; OR, odds ratio; CI, confidence interval; P, p-value.

**Supplementary Table 5**: CCSS replication results for top genetic associations with self-reported insulin-dependent diabetes mellitus risk (European genetic ancestry)

| | | | | CCSS Original (N=108 cases) | | | | CCSS Expansion (N=25 cases) | | | |
|---|---|---|---|---|---|---|---|---|---|---|---|
| Chr | BP | EA | NEA | EAF | OR | CI | P | EAF | OR | CI | P |
| 2 | 2460867 | C | T | 0.14 | 1.45 | 1.00-2.10 | 0.047 | 0.13 | 0.91 | 0.38-2.14 | 0.82 |
| 2 | 2462940 | A | G | 0.14 | 1.49 | 1.03-2.14 | 0.033 | 0.13 | 0.91 | 0.38-2.14 | 0.82 |
| 8 | 50635641 | T | C | 0.16 | 1.56 | 1.12-2.17 | $7.9 \times 10^{-3}$ | 0.15 | 0.52 | 0.23-1.20 | 0.12 |
| 8 | 50636999 | C | T | 0.16 | 1.56 | 1.12-2.17 | $8.1 \times 10^{-3}$ | 0.15 | 0.52 | 0.22-1.20 | 0.13 |
| 8 | 50637285 | A | T | 0.16 | 1.56 | 1.12-2.17 | $8.1 \times 10^{-3}$ | 0.15 | 0.52 | 0.22-1.20 | 0.13 |
| 8 | 50637444 | A | G | 0.16 | 1.56 | 1.12-2.17 | $8.1 \times 10^{-3}$ | 0.15 | 0.52 | 0.22-1.20 | 0.13 |
| 8 | 50638825 | G | A | 0.16 | 1.56 | 1.12-2.17 | $8.1 \times 10^{-3}$ | 0.15 | 0.52 | 0.22-1.20 | 0.13 |
| 8 | 50641552 | A | G | 0.16 | 1.56 | 1.12-2.17 | $8.1 \times 10^{-3}$ | 0.15 | 0.52 | 0.22-1.20 | 0.13 |
| 8 | 50643118 | T | G | 0.16 | 1.56 | 1.12-2.17 | $8.1 \times 10^{-3}$ | 0.15 | 0.52 | 0.22-1.20 | 0.13 |
| 8 | 50644980 | A | G | 0.18 | 1.43 | 1.04-1.99 | 0.029 | 0.18 | 0.83 | 0.37-1.83 | 0.64 |
| 8 | 50661918 | G | C | 0.69 | 0.88 | 0.66-1.17 | 0.36 | 0.70 | 1.09 | 0.61-1.96 | 0.77 |
| 8 | 50666979 | A | G | 0.22 | 1.08 | 0.79-1.49 | 0.63 | 0.22 | 1.08 | 0.54-2.15 | 0.82 |
| 8 | 50667110 | A | G | 0.22 | 1.1 | 0.80-1.51 | 0.55 | 0.22 | 1.08 | 0.54-2.16 | 0.82 |
| 8 | 50667850 | G | T | 0.22 | 1.1 | 0.80-1.51 | 0.55 | 0.22 | 1.08 | 0.55-2.16 | 0.82 |
| 8 | 50672410 | T | A | 0.22 | 1.1 | 0.80-1.51 | 0.55 | 0.22 | 1.08 | 0.54-2.16 | 0.82 |
| 8 | 50672830 | C | T | 0.22 | 1.09 | 0.79-1.49 | 0.61 | 0.21 | 1.09 | 0.55-2.16 | 0.82 |
| 8 | 50673224 | G | A | 0.22 | 1.1 | 0.80-1.51 | 0.55 | 0.22 | 1.08 | 0.54-2.16 | 0.82 |
| 8 | 50674706 | G | A | 0.22 | 1.1 | 0.80-1.51 | 0.55 | 0.22 | 1.08 | 0.54-2.15 | 0.82 |
| 8 | 50675283 | G | A | 0.22 | 1.1 | 0.80-1.51 | 0.55 | 0.22 | 1.08 | 0.54-2.16 | 0.82 |
| 8 | 50676103 | C | T | 0.22 | 1.06 | 0.77-1.46 | 0.71 | 0.21 | 1.09 | 0.55-2.17 | 0.81 |
| 8 | 50676751 | G | T | 0.22 | 1.07 | 0.77-1.47 | 0.70 | 0.21 | 1.09 | 0.55-2.17 | 0.81 |
| 8 | 50681567 | G | A | 0.21 | 1.1 | 0.80-1.52 | 0.56 | 0.20 | 1.17 | 0.59-2.31 | 0.66 |
| 19 | 22614854 | G | A | 0.06 | 1.04 | 0.57-1.89 | 0.91 | 0.06 | 0.75 | 0.22-2.60 | 0.65 |
| 19 | 22614973 | A | G | 0.06 | 1.04 | 0.57-1.89 | 0.90 | 0.06 | 0.77 | 0.22-2.67 | 0.68 |
| 19 | 22615242 | T | C | 0.06 | 1.04 | 0.57-1.89 | 0.91 | 0.06 | 0.75 | 0.22-2.60 | 0.65 |
| 19 | 22615378 | A | G | 0.06 | 1.04 | 0.57-1.89 | 0.91 | 0.06 | 0.75 | 0.22-2.60 | 0.65 |
| 19 | 22615432 | C | A | 0.06 | 1.04 | 0.57-1.89 | 0.91 | 0.06 | 0.76 | 0.22-2.61 | 0.66 |

Abbreviations: CCSS, Childhood Cancer Survivor Study; Chr, chromosome; BP, base position, GRCh38 (hg38) build; EA, effect (risk) allele; NEA, non-effect (reference) allele; OR, odds ratio; CI, confidence interval; P, p-value.

**Supplementary Table 6**: Associations between index variants and self-reported and diagnosed type 2 diabetes risk in the general population (UK Biobank Study)

| | Self-reported T2D (2,292 cases) | | | | | |
|---|---|---|---|---|---|---|
| Variants | Minor allele | UKB MAF | N | Beta | SE | P |
| 2:2438081:AAAATAAAATAAAATAAAATAAAAAC:A | A | 0.20 | 361141 | $2.4 \times 10^{-4}$ | $2.4 \times 10^{-4}$ | 0.32 |
| 8:51551385:A:G | G | 0.15 | 361141 | $9.0 \times 10^{-5}$ | $2.6 \times 10^{-4}$ | 0.73 |
| 19:22797656:A:G | G | 0.06 | 361141 | $1.1 \times 10^{-3}$ | $4.1 \times 10^{-4}$ | $6.6 \times 10^{-3}$ |
| 5:9843018:T:C | Not present | | | | | |
| Variants | ICD T2D (888 cases) | | | | | |
| 2:2438081:AAAATAAAATAAAATAAAATAAAAAC:A | A | 0.20 | 361194 | $4.4 \times 10^{-4}$ | $1.5 \times 10^{-4}$ | $3.6 \times 10^{-3}$ |
| 8:51551385:A:G | G | 0.15 | 361194 | $2.5 \times 10^{-4}$ | $1.6 \times 10^{-4}$ | 0.12 |
| 19:22797656:A:G | G | 0.06 | 361194 | $3.4 \times 10^{-4}$ | $2.6 \times 10^{-4}$ | 0.18 |
| 5:9843018:T:C | Not present | | | | | |

Variants included in genes are annotated in chromosome:base position:non-effect allele:effect allele format, GRCh37 build. Self-reported T2D summary statistics were obtained from file 20002_1223.gwas.imputed_v3.both_sexes.tsv.bgz and ICD-10-coded T2D summary statistics were obtained from file E4_DM2.gwas.imputed_v3.both_sexes.tsv.bgz**.**
Abbreviations: UKB, UK Biobank Study; MAF, minor allele frequency; SE, standard error; P, p-value.

**Supplementary Table 7**: Abdominal radiation therapy-stratified analysis of diabetes mellitus risk index SNPs in SJLIFE survivors of European and African ancestry

| | Chr | BP | EA | NEA | Stratification | N | OR | CI | P |
|---|---|---|---|---|---|---|---|---|---|
| SJLIFE EUR | 2 | 2434309 | A | AAAATAAAATAAA ATAAAATAAAAAC | All survivors | 3073 | 1.99 | 1.49-2.66 | $2.8x10^{-6}$ |
| | | | | | No abdominal RT | 2512 | 2.07 | 1.48-2.89 | $2.3x10^{-5}$ |
| | | | | | Any abdominal RT | 561 | 1.79 | 1.00-3.23 | 0.052 |
| | 5 | 9843018 | C | T | All survivors | 3102 | 2.47 | 1.23-4.98 | 0.011 |
| | | | | | No abdominal RT | 2536 | 2.55 | 1.10-5.94 | 0.029 |
| | | | | | Any abdominal RT | 566 | 2.01 | 0.56-7.24 | 0.29 |
| | 8 | 50638825 | G | A | All survivors | 3100 | 1.99 | 1.56-2.55 | $4.4x10^{-8}$ |
| | | | | | No abdominal RT | 2535 | 2.17 | 1.63-2.90 | $1.4x10^{-7}$ |
| | | | | | Any abdominal RT | 565 | 1.63 | 0.99-2.68 | 0.055 |
| | 19 | 22614854 | G | A | All survivors | 3100 | 2.26 | 1.60-3.20 | $4.1x10^{-6}$ |
| | | | | | No abdominal RT | 2534 | 2.24 | 1.47-3.41 | $1.8x10^{-4}$ |
| | | | | | Any abdominal RT | 566 | 2.37 | 1.23-4.57 | 0.010 |
| SJLIFE AFR | Chr | BP | EA | NEA | Stratification | N | OR | CI | P |
| | 2 | 2434309 | A | AAAATAAAATAAA ATAAAATAAAAAC | All survivors | 556 | 2.30 | 1.30-4.07 | $4.2x10^{-3}$ |
| | | | | | No abdominal RT | 454 | 2.69 | 1.35-5.39 | $5.1x10^{-3}$ |
| | | | | | Any abdominal RT | 102 | 12.10 | 0.98-148.94 | 0.052 |
| | 5 | 9843018 | C | T | All survivors | 574 | 10.19 | 4.12-25.21 | $5.1x10^{-7}$ |
| | | | | | No abdominal RT | 468 | 12.95 | 4.09-40.97 | $1.3x10^{-5}$ |
| | | | | | Any abdominal RT | 106 | 90.85 | 2.25-3660.27 | 0.017 |
| | 8 | 50638825 | G | A | All survivors | 573 | 0.53 | 0.30-0.91 | 0.022 |
| | | | | | No abdominal RT | 467 | 0.38 | 0.17-0.83 | 0.016 |
| | | | | | Any abdominal RT | 106 | 0.44 | 0.14-1.41 | 0.17 |
| | 19 | 22614854 | G | A | All survivors | 573 | 2.99 | 1.66-5.37 | $2.6x10^{-4}$ |
| | | | | | No abdominal RT | 467 | 2.72 | 1.28-5.78 | $9.6x10^{-3}$ |
| | | | | | Any abdominal RT | 106 | 5.15 | 1.08-24.60 | 0.040 |

Abbreviations: SJLIFE, St. Jude Lifetime Cohort Study; Chr, chromosome; BP, base position, GRCh38 (hg38) build; EA, effect (risk) allele; NEA, non-effect (reference) allele; EUR, European genetic ancestry; AFR, African genetic ancestry; RT, radiation therapy; OR, odds ratio; CI, confidence interval; P, p-value.

**Supplementary Table 8**: Alkylating agent-stratified analysis of diabetes mellitus risk index SNPs in SJLIFE survivors of European and African ancestry

| | Chr | BP | EA | NEA | Stratification | N | OR | CI | P |
|---|---|---|---|---|---|---|---|---|---|
| SJLIFE EUR | 2 | 2434309 | A | AAAATAAAATAAAATAAAATAAAAAC | All survivors | 3073 | 1.99 | 1.49-2.66 | $2.8x10^{-6}$ |
| | | | | | No abdominal RT, no AA | 1154 | 1.87 | 1.07-3.27 | 0.027 |
| | | | | | No abdominal RT, high AA (≥4000 mg/m$^2$) | 1046 | 2.37 | 1.45-3.87 | $6.0x10^{-4}$ |
| | 5 | 9843018 | C | T | All survivors | 3102 | 2.47 | 1.23-4.98 | 0.011 |
| | | | | | No AA | 1351 | 1.27 | 0.32-5.00 | 0.73 |
| | | | | | Any AA | 1751 | 3.32 | 1.44-7.68 | $5.0x10^{-3}$ |
| | 8 | 50638825 | G | A | All survivors | 3100 | 1.99 | 1.56-2.55 | $4.4x10^{-8}$ |
| | | | | | No abdominal RT, no AA | 1168 | 3.17 | 1.96-5.12 | $2.5x10^{-6}$ |
| | | | | | No abdominal RT, high AA (≥4000 mg/m$^2$) | 1052 | 1.57 | 1.03-2.40 | 0.038 |
| | 19 | 22614854 | G | A | All survivors | 3100 | 2.26 | 1.60-3.20 | $4.1x10^{-6}$ |
| | | | | | No abdominal RT, no AA | 1169 | 1.63 | 0.74-3.57 | 0.22 |
| | | | | | No abdominal RT, high AA (≥4000 mg/m$^2$) | 1051 | 3.32 | 1.91-5.79 | $2.3x10^{-5}$ |
| SJLIFE AFR | Chr | BP | EA | NEA | Stratification | N | OR | CI | P |
| | 2 | 2434309 | A | AAAATAAAATAAAATAAAATAAAAAC | All survivors | 556 | 2.30 | 1.30-4.07 | $4.2x10^{-3}$ |
| | | | | | No AA | 267 | 2.39 | 0.99-5.76 | 0.053 |
| | | | | | High AA (≥4000 mg/m$^2$) | 215 | 3.95 | 1.38-11.31 | 0.010 |
| | 5 | 9843018 | C | T | All survivors | 574 | 10.19 | 4.12-25.21 | $5.1x10^{-7}$ |
| | | | | | No AA | 275 | 10.84 | 3.08-38.18 | $2.1x10^{-4}$ |
| | | | | | Any AA | 299 | 17.81 | 3.46-91.60 | $5.7x10^{-4}$ |
| | 8 | 50638825 | G | A | All survivors | 573 | 0.53 | 0.30-0.91 | 0.022 |
| | | | | | No AA | 274 | 0.62 | 0.29-1.34 | 0.23 |
| | | | | | High AA (≥4000 mg/m$^2$) | 221 | 0.47 | 0.18-1.19 | 0.11 |
| | 19 | 22614854 | G | A | All survivors | 573 | 2.99 | 1.66-5.37 | $2.6x10^{-4}$ |
| | | | | | No AA | 275 | 2.00 | 0.88-4.54 | 0.096 |
| | | | | | High AA (≥4000 mg/m$^2$) | 220 | 5.74 | 1.99-16.54 | $1.2x10^{-3}$ |

Abbreviations: SJLIFE, St. Jude Lifetime Cohort Study; Chr, chromosome; BP, base position, GRCh38 (hg38) build; EA, effect (risk) allele; NEA, non-effect (reference) allele; EUR, European genetic ancestry; AFR, African genetic ancestry; AA, alkylating agents; RT, radiation therapy; m, meters; mg, milligrams, OR, odds ratio; CI, confidence interval; P, p-value.

**Supplementary Table 9**: BMI-stratified analysis of the late diabetes risk index SNP in the chromosome 8 locus among SJLIFE and CCSS survivors

| Chr | BP | EA | NEA | Study sample | BMI stratification | N | OR | 95% CI | P |
|---|---|---|---|---|---|---|---|---|---|
| 8 | 50638825 | G | A | SJLIFE EUR | BMI>25 kg/m$^2$ | 1941 | 2.23 | 1.72-2.89 | 1.8x10$^{-9}$ |
| | | | | | BMI≤25 kg/m$^2$ | 1159 | 0.90 | 0.42-1.94 | 0.80 |
| | | | | CCSS ORIGINAL (EUR) | BMI>25 kg/m$^2$ | 2202 | 1.69 | 1.15-2.47 | 7.0x10$^{-3}$ |
| | | | | | BMI≤25 kg/m$^2$ | 1585 | 1.18 | 0.60-2.29 | 0.63 |

Abbreviations: SJLIFE, St. Jude Lifetime Cohort Study; CCSS, Childhood Cancer Survivor Study; Chr, chromosome; BP, base position, GRCh38 (hg38) build; EA, effect (risk) allele; NEA, non-effect (reference) allele; EUR, European genetic ancestry; BMI, body mass index; m, meters; kg, kilograms, OR, odds ratio; CI, confidence interval; P, p-value.

**Supplementary Table 10**: Multi-ancestry T2D PRS (Mahajan et al.) quintile associations with diabetes mellitus risk in survivors

| Cohort | Treatment | N | Q | OR (95% CI) | P | Treatment | N | Q | OR (95% CI) | P |
|---|---|---|---|---|---|---|---|---|---|---|
| SJLIFE AFR | No AA | 275 | Q2 | 5.68 (0.82-39.25) | 0.078 | No abd RT | 468 | Q2 | 14.82 (1.49-147.11) | 0.021 |
| | | | Q3 | 1.19 (0.14-9.94) | 0.87 | | | Q3 | 4.43 (0.42-46.50) | 0.21 |
| | | | Q4 | 4.51 (0.68-29.77) | 0.12 | | | Q4 | 13.48 (1.32-137.80) | 0.028 |
| | | | Q5 | 4.48 (0.72-27.76) | 0.11 | | | Q5 | 18.49 (1.88-181.58) | 0.012 |
| | Any AA | 299 | Q2 | 4.30 (0.37-49.84) | 0.24 | Any abd RT | 106 | Q2 | 0.70 (0.03-18.68) | 0.83 |
| | | | Q3 | 3.52 (0.31-40.51) | 0.31 | | | Q3 | 1.29 (0.08-20.67) | 0.86 |
| | | | Q4 | 3.14 (0.24-41.70) | 0.39 | | | Q4 | 0.04 (0.00-8.94) | 0.24 |
| | | | Q5 | 13.85 (1.24-154.48) | 0.033 | | | Q5 | 4.28 (0.40-46.27) | 0.23 |
| Combined EUR | No AA, no abd RT | 3561 | Q2 | 1.59 (0.77-3.30) | 0.21 | No abd RT | 7171 | Q2 | 1.66 (1.02-2.70) | 0.04 |
| | | | Q3 | 2.61 (1.34-5.10) | 4.9x10$^{-3}$ | | | Q3 | 2.46 (1.56-3.89) | 1.1x10$^{-4}$ |
| | | | Q4 | 2.02 (1.00-4.05) | 0.048 | | | Q4 | 2.49 (1.57-3.94) | 1.0x10$^{-4}$ |
| | | | Q5 | 4.07 (2.16-7.69) | 1.5x10$^{-5}$ | | | Q5 | 4.90 (3.19-7.52) | 3.6x10$^{-13}$ |
| | Any AA, no abd RT | 3610 | Q2 | 1.66 (0.86-3.22) | 0.13 | Any abd RT | 1896 | Q2 | 0.95 (0.49-1.83) | 0.87 |
| | | | Q3 | 2.32 (1.23-4.38) | 9.3x10$^{-3}$ | | | Q3 | 1.96 (1.09-3.54) | 0.025 |
| | | | Q4 | 2.84 (1.52-5.29) | 1.0x10$^{-3}$ | | | Q4 | 2.67 (1.52-4.71) | 6.7x10$^{-4}$ |
| | | | Q5 | 5.86 (3.25-10.57) | 4.4x10$^{-9}$ | | | Q5 | 3.62 (2.08-6.30) | 5.0x10$^{-6}$ |
| | Higher-dose AA, no abd RT | 2695 | Q2 | 2.00 (0.88-4.53) | 0.099 | Higher-dose abd RT | 1431 | Q2 | 0.94 (0.47-1.90) | 0.87 |
| | | | Q3 | 2.82 (1.29-6.18) | 9.3x10$^{-3}$ | | | Q3 | 2.00 (1.07-3.76) | 0.031 |
| | | | Q4 | 3.44 (1.59-7.42) | 1.7x10$^{-3}$ | | | Q4 | 2.08 (1.11-3.88) | 0.022 |
| | | | Q5 | 8.43 (4.06-17.53) | 1.1x10$^{-8}$ | | | Q5 | 3.56 (1.97-6.43) | 2.5x10$^{-5}$ |
| SJLIFE EUR | No AA, no abd RT | 1169 | Q2 | 1.42 (0.50-4.00) | 0.51 | No abd RT | 2536 | Q2 | 1.46 (0.75-2.81) | 0.26 |
| | | | Q3 | 2.11 (0.81-5.51) | 0.13 | | | Q3 | 1.87 (1.00-3.52) | 0.051 |
| | | | Q4 | 1.55 (0.56-4.31) | 0.4 | | | Q4 | 2.36 (1.28-4.36) | 5.9x10$^{-3}$ |
| | | | Q5 | 3.74 (1.51-9.25) | 4.4x10$^{-3}$ | | | Q5 | 4.67 (2.62-8.32) | 1.7x10$^{-7}$ |
| | Any AA, no abd RT | 1367 | Q2 | 1.47 (0.61-3.51) | 0.39 | Any abd RT | 566 | Q2 | 1.73 (0.59-5.01) | 0.32 |
| | | | Q3 | 1.69 (0.72-3.98) | 0.23 | | | Q3 | 3.69 (1.33-10.25) | 0.012 |
| | | | Q4 | 2.97 (1.34-6.56) | 7.3x10$^{-3}$ | | | Q4 | 4.20 (1.54-11.47) | 5.2x10$^{-3}$ |
| | | | Q5 | 5.62 (2.60-12.15) | 1.1x10$^{-5}$ | | | Q5 | 6.44 (2.37-17.47) | 2.6x10$^{-4}$ |
| | Higher-dose AA, no abd RT | 1052 | Q2 | 1.86 (0.64-5.41) | 0.25 | Higher-dose abd RT | 427 | Q2 | 2.50 (0.75-8.31) | 0.13 |
| | | | Q3 | 2.33 (0.82-6.58) | 0.11 | | | Q3 | 4.57 (1.40-14.92) | 0.012 |
| | | | Q4 | 3.50 (1.33-9.26) | 0.011 | | | Q4 | 5.38 (1.66-17.44) | 5.0x10$^{-3}$ |
| | | | Q5 | 8.83 (3.41-22.84) | 7.0x10$^{-6}$ | | | Q5 | 7.92 (2.50-25.08) | 4.4x10$^{-4}$ |
| CCSS EUR | No AA, no abd RT | 2392 | Q2 | 1.71 (0.60-4.89) | 0.32 | No abd RT | 4635 | Q2 | 1.69 (0.81-3.52) | 0.16 |
| | | | Q3 | 3.06 (1.18-7.99) | 0.022 | | | Q3 | 2.93 (1.49-5.73) | 1.8x10$^{-3}$ |
| | | | Q4 | 2.43 (0.91-6.49) | 0.077 | | | Q4 | 2.35 (1.17-4.73) | 0.016 |
| | | | Q5 | 4.42 (1.77-11.08) | 1.5x10$^{-3}$ | | | Q5 | 5.01 (2.63-9.54) | 9.3x10$^{-7}$ |
| | Any AA, no abd RT | 2243 | Q2 | 1.64 (0.58-4.63) | 0.35 | Any abd RT | 1330 | Q2 | 0.52 (0.21-1.25) | 0.14 |
| | | | Q3 | 2.92 (1.12-7.64) | 0.029 | | | Q3 | 1.27 (0.61-2.66) | 0.52 |
| | | | Q4 | 2.31 (0.85-6.31) | 0.10 | | | Q4 | 1.94 (0.97-3.88) | 0.059 |
| | | | Q5 | 5.85 (2.32-14.73) | 1.8x10$^{-4}$ | | | Q5 | 2.46 (1.26-4.81) | 8.6x10$^{-3}$ |
| | Higher-dose AA, no abd RT | 1643 | Q2 | 1.72 (0.48-6.21) | 0.41 | Higher-dose abd RT | 898 | Q2 | 0.40 (0.15-1.07) | 0.069 |
| | | | Q3 | 3.06 (0.94-9.92) | 0.062 | | | Q3 | 1.19 (0.53-2.70) | 0.67 |
| | | | Q4 | 2.48 (0.72-8.54) | 0.15 | | | Q4 | 1.14 (0.51-2.55) | 0.74 |
| | | | Q5 | 7.23 (2.35-22.25) | 5.6x10$^{-4}$ | | | Q5 | 2.18 (1.05-4.54) | 0.036 |

Abbreviations: SJLIFE, St. Jude Lifetime Cohort Study; CCSS, Childhood Cancer Survivor Study; EUR, European genetic ancestry; AFR, African genetic ancestry; AA, alkylating agents; abd RT, abdominal radiation therapy; Q, PRS quintile; OR, odds ratio; CI, confidence interval; P, p-value.
Higher dose AA defined by doses ≥4,000 mg/m$^2$; higher doses of abd RT defined by doses ≥2,000 cGy.

**Supplementary Table 11:** Top genes enriched for rare pathogenic variants associated with diabetes mellitus risk in SJLIFE childhood cancer survivors of European genetic ancestry ($P<1x10^{-3}$)

| | | Discovery (N=1550; 127 cases) | Replication (N=1552; 134 cases) |
|---|---|---|---|
| Gene | Genetic variants | Burden test P | Burden test P |
| ENSG00000254469 (*XNDC1N*) | chr11:71918924:G:A; chr11:71918995:TTTAGCAAGTTCTCTACAGGA:T | $8.5x10^{-4}$ | $9.4x10^{-3}$ |
| *ZNF655* | chr7:99563862:G:A; chr7:99563873:CCGCCTTCCCA:C | $7.1x10^{-4}$ | 0.21 |

Variants included in genes are annotated in chromosome:base position:non-effect allele:effect allele format, GRCh38 build.

**Supplementary Table 12:** Single variant association test results for *XNDC1N* rare pathogenic/likely pathogenic variants

| | Combined, SJLIFE European genetic ancestry (N=3102) | | |
|---|---|---|---|
| Rare variant association test type | MAF (MAC) | OR (95% CI) | P |
| Single variant: chr11:71918924:G:A | 0.002 (12) | 6.30 (1.60-24.86) | $8.6x10^{-3}$ |
| Single variant: chr11:71918995:TTTAGCAAGTTCTCTACAGGA:T | 0.001 (9) | 13.42 (2.77-64.99) | $1.2x10^{-3}$ |

Abbreviations: MAF(C), minor allele frequency (count); OR, odds ratio; CI, confidence interval; P, p-value.
Variants are annotated in chromosome:base position:non-effect allele:effect allele format, GRCh38 build.

**Supplementary Table 13**: Comparing *XNDC1N* rare pathogenic risk allele frequencies among healthy adults (gnomAD, N=16,465) and SJLIFE survivors without diabetes mellitus (N=2,841) and with diabetes mellitus (N=261)

| *XNDC1N* rare pathogenic variants (rsid) | Study sample | MAF | Allele count | Allele number | OR (95% CI) | P |
|---|---|---|---|---|---|---|
| chr11:71918924:G:A (rs200142185) | gnomAD: EUR controls | 0.0020 | 14 | 6850 | Reference | |
| | EUR survivors without DM | 0.0012 | 7 | 5704 | 0.60 (0.21-1.60) | 0.38 |
| | EUR survivors with DM | 0.0096 | 5 | 522 | 4.72 (1.33-13.94) | $8.8 \times 10^{-3}$ |
| chr11:71918995:TTTAGCAAGTTCTCTACAGGA:T (rs771623613) | gnomAD: EUR controls | 0.0003 | 2 | 6848 | Reference | |
| | EUR survivors without DM | 0.0009 | 5 | 5682 | 3.01 (0.49-31.66) | 0.26 |
| | EUR survivors with DM | 0.0057 | 3 | 522 | 19.77 (2.26-236.43) | $3.2 \times 10^{-3}$ |

Abbreviations: Genome Aggregation Database, gnomAD; EUR, European genetic ancestry; DM, diabetes mellitus; rsid, genetic variant identifier using dbSNP build 151; MAF, minor allele frequency; OR, odds ratio; CI, confidence interval; P, p-value. Variants are also shown in chr:base position:reference allele:effect allele format, where base position is provided in GRCh38 (hg38) build. P-values are from 2-sided Fisher's exact tests comparing allele counts, using the European (non-Finnish) control/biobank samples in gnomAD v3.1.2.

**Supplementary Table 14**: Index variant *in silico* functional annotations

| rsid | Chr | BP | EA | NEA | Genes within 5 kb (closest) | Functional consequence | CADD | Regulatory function | meQTLs | eQTLs/sQTLs |
|---|---|---|---|---|---|---|---|---|---|---|
| rs143578296 | 2 | 2434309 | A | AAAATAAAATAAAA TAAAATAAAAAC | | | 6.5 | | Not available | Not available |
| rs13436710 | 5 | 9843018 | C | T | *LINC02112* | Intron variant | 3.3 | | Not available | Single-tissue sQTLs: *FAM173B* (brain-hypothalamus, EAdir=+) |
| rs6993752 | 8 | 50638825 | G | A | *SNTG1* | Intron variant | 0.1 | CTCF binding site (ENSR00000854839); enhancer (ENSR00001137992) | *SNTG1* (EAdir=+, Probe=cg17991843) | |
| rs200142185 | 11 | 71918924 | A | G | *XNDC1N* | Incomplete terminal codon variant, coding sequence variant; stop gained | 40.0 | | Not available | Not available |
| rs771623613 | 11 | 71918995 | T | TTTAGCAAG TTCTCTACAGGA | *XNDC1N* | Stop gained, frameshift variant | 33.0 | | Not available | Not available |
| rs35017858 | 19 | 22614854 | G | A | *LINC01785* | Downstream gene variant | 5.0 | | Not available | Single-tissue eQTLs: *LINC01785* (thyroid, EAdir=-) ZNF723P (testis, EAdir=+) *ZNF257* (brain nucleus accumbens basal ganglia, EAdir=-) *ZNF492* (esophagus mucosa, cells cultured fibroblasts, testis, EAdir=-) |

Major abbreviations: rsid, genetic variant identifier, dbSNP build 151; Chr, chromosome; BP: genomic base position, GRCh38 build; EA, effect allele; NEA, non-effect allele; QTL, quantitative trait loci; eQTL, expression QTL; sQTL, splicing QTL; meQTL methylation QTL.

a. Functional consequences were annotated with Ensembl Variant Effect Predictor (VEP v99).
b. Combined Annotation Dependent Depletion (CADD) scores reflect variant deleteriousness, PHRED-scaled such that scores >10 represent variants with the top 10% of CADD scores, >20 with top 1% of CADD scores, etc.
c. Single-tissue eQTLs/sQTLs with FDR≤5% were based on BIOS QTL, eQTLGen, and GTEx v8, where top eQTL/sQTL genes are genes with at least one significant (FDR<5%) *cis*-SNP association.
d. BIOS QTL was used to annotate significant (FDR<5%) meQTL variants.

**Supplementary Table 15**: Chromatin state annotations in pancreas-, liver-, and fat-related epigenomes

| | | | Any promoter | | Any enhancer | | Any transcription | | Any polycomb repressed | |
|---|---|---|---|---|---|---|---|---|---|---|
| rsid | CHR | BP | Overlapping variants | Relevant epigenomes | Overlapping variants | Relevant epigenomes | Overlapping variants | Relevant epigenomes | Overlapping variants | Relevant epigenomes |
| rs143578296 | 2 | 2434309 | | | | | | | **chr2:2438081**, chr2:2464639, chr2:2466712 | LIVER (E066,E118); PANCREAS (E087) |
| rs13436710 | 5 | 9843018 | | | | | | | **chr5:9843130**, chr5:9853144, chr5:9841295 | FAT (E063); LIVER (E066); PANCREAS (E087) |
| rs6993752 | 8 | 50638825 | chr8:51555678, chr8:51554112 | PANCREAS (E087) | chr8:51595171, chr8:51767887, chr8:51588663, chr8:51554112, chr8:51555678, chr8:51631199, chr8:51634561, chr8:51631400, chr8:51627432, chr8:51627574, **chr8:51551385**, chr8:51557540, chr8:51807451, chr8:51729352 | LIVER (E066); PANCREAS (E087) | | | | |
| rs35017858 | 19 | 22614854 | | | | | | | chr19:22795650, chr19:22795896, **chr19:22797656**, chr19:22797775, chr19:22798044, chr19:22798180, chr19:22798234 | PANCREAS (E087) |
| rs200142185 | 11 | 71918924 | | | | | **chr11:71629970, chr11:71630041** | FAT (E023,E025,E063); LIVER (E066,E118); PANCREAS (E098) | | |
| rs771623613 | 11 | 71918995 | | | | | | | | |

Chromatin state annotations were taken from the 15-state (ChromHMM) model based on 12 epigenetic marks for 127 epigenomes (Roadmap Epigenomics Consortium). We used pooled chromatin state annotations for any promoter (states 1-2, 10), transcription (states 3-5), enhancer (states 6-7, 11-12), and polycomb repressed (states 13-14) states among relevant epigenomes (liver [E066, E118]; pancreas [E087, E098]; fat [E023, E025, E063]). Overlapping variants are annotated in Chr:BP format, GRCh37 build; bolded variants correspond to index variants.
Major abbreviations: rsid, genetic variant identifier, dbSNP build 151; Chr, chromosome; BP: genomic base position, GRCh38 build.
